# BOLD: Blood-gas and Oximetry Linked Dataset – Open Source Research

**DOI:** 10.1101/2023.10.03.23296485

**Authors:** João Matos, Tristan Struja, Jack Gallifant, Luis Nakayama, Marie-Laure Charpignon, Xiaoli Liu, Nicoleta Economou-Zavlanos, Jaime S. Cardoso, Kimberly S Johnson, Nrupen Bhavsar, Judy Gichoya, Leo Anthony Celi, A. Ian Wong

## Abstract

Pulse oximeters measure peripheral arterial oxygen saturation (SpO_2_) noninvasively, while the gold standard (SaO_2_) involves arterial blood gas measurement. There are known racial and ethnic disparities in their performance. BOLD is a new comprehensive dataset that aims to underscore the importance of addressing biases in pulse oximetry accuracy, which disproportionately affect darker-skinned patients.

The dataset was created by harmonizing three Electronic Health Record databases (MIMIC-III, MIMIC-IV, eICU-CRD) comprising Intensive Care Unit stays of US patients. Paired SpO_2_ and SaO_2_ measurements were time-aligned and combined with various other sociodemographic and parameters to provide a detailed representation of each patient. BOLD includes 49,099 paired measurements, within a 5-minute window and with oxygen saturation levels between 70-100%. Minority racial and ethnic groups account for ∼25% of the data – a proportion seldom achieved in previous studies. The codebase is publicly available.

Given the prevalent use of pulse oximeters in the hospital and at home, we hope that BOLD will be leveraged to develop debiasing algorithms that can result in more equitable healthcare solutions.

## Background & Summary

The measurement and management of arterial blood gas (ABG) and pulse oximetry in the Intensive Care Unit (ICU) have long been the subject of clinical interest but are often under-studied. Pulse oximeters and ABG are tools for evaluating systemic oxygen saturation and providing guidance for clinical decision-making. Standardization in pairing arterial blood gas samples with pulse oximeter readings, a critical component for effective patient monitoring and management, is particularly scarce. This is due in part to challenges in coordinating large electronic health record (EHR) datasets and synchronizing clinical protocols across multiple medical centers.

Recent research by Sjoding et al., Wong et al., Valbuena et al., and Gottlieb et al., has added another layer of complexity by uncovering racial disparities in pulse oximeter reading accuracy, which have critical implications for patient care and outcomes.^1–4^ Such disparities further emphasize the urgent need for robust and inclusive datasets allowing the conduct of thorough comparative analysis across subpopulations. Given these pervasive challenges and recent findings, the retrospective investigation of real-world data can offer invaluable insights. All of the above-listed studies have used EHR data, which was stored in multiple formats that may not be easy to use and may not be available to external researchers, representing a significant barrier to entry.

Existing large-scale EHR datasets, even when available in open access or under a formal research protocol, are often not in a form that can help readily answer nuanced yet urgent clinical questions such as the need of applying potential corrections by skin tone to the measurements output by FDA-approved devices.^5–7^ The understanding of health systems, data schemas, and critical care physiology necessary to make individual ICU-EHR datasets usable in practice is nontrivial; thus, our effort to preprocess raw time series to create a unified dataset removes a barrier to entry.

This paper aims to present a comprehensive approach to the extraction, processing, and analysis of arterial blood gas samples and pulse oximeter readings from electronic health records (EHR). We demonstrate the application of this principled approach to the issue of racial and ethnic disparities in device measurement and hope it can guide future studies.

We propose a clinically-grounded and reproducible methodology to convert unprocessed database queries into a clinically useful dataset. Our multidisciplinary team, which includes clinicians (i.e., pulmonologists and intensivists) and data scientists, has developed rules based on clinical and physiological standards for pairing each arterial blood gas sample with a corresponding pulse oximeter reading as well as with clinical scores, vital signs, and laboratory test values.

Our primary objective is to facilitate extensive analysis of pulse oximetry by merging data from three major, publicly available, ICU-EHR databases – MIMIC-III, MIMIC-IV, and eICU-CRD. A combined dataset not only offers a solution to the paucity of large and diverse datasets but also a unique platform for identifying disparities in pulse oximetry readings and designing approaches to remediate such inequities. By making this robust dataset publicly available, we provide researchers with the means to develop models that address known racial and ethnic disparities and those yet to be discovered, with the potential to improve fairness in healthcare delivery. Furthermore, by making the platform and code available, this platform can serve as an example for conducting similar studies that would benefit from linked databases.

Our work stands as a necessary and fundamental milestone to any machine learning (ML) ^8^ or advanced data analysis of oxygen readings that can be performed to produce actionable insights. Data curation is a critical step, especially considering the volume of data in our base datasets; e.g., MIMIC-III v.1.4 alone contains over 58,000 hospital admissions from approximately 38,600 adults, resulting in 6.2GB of data. A traditional manual chart review (e.g., to identify patients at risk of hypoxemia) would be impractical given this volume of data, emphasizing the need for an automated, yet clinically-validated approach. As an example, we anticipate value in a machine learning model that, based on a patient’s oxygen saturation trajectory since ICU admission, could predict the likelihood of hypoxemia in the next hours.

The dataset we present not only addresses the critical issue of pulse oximetry disparities but also offers a versatile tool for the broader medical research community. In the future, we plan to extend this dataset to other EHR databases and to include waveform data. By detailing our methodologies and sharing our modular scripts, we provide avenues for other researchers to build upon this work, potentially extending it to other biometric readings (e.g., body temperature, blood pressure) and clinical contexts (e.g., home-based care, primary care, emergency room).

Overall, our dataset aims to serve as a pivotal resource for the clinical and research communities alike, informing respiratory parameter management in the ICU with a particular focus on addressing racial and ethnic disparities in pulse oximetry accuracy. We operationalize the evaluation of racial and ethnic disparities in pulse oximetry by quantifying differences in the occurrence of hidden hypoxemia, defined as SaO2 > 88% but SpO_2_ ≥ 88%. ^2^ Finally, we provide the complete codebases for data curation and validation assays to encourage ongoing, collaborative research in this critical area.

As observational data collected in hospital settings are often used retrospectively to inform the development, manufacturing, and quality control of pulse oximeters, our effort should prompt other parties (e.g., pulse oximetry equipment manufacturers) with access to such paired measurements but in different settings (e.g., randomized trials) to also share the underlying datasets with the public.

## Methods

### Data sources

Three EHR databases were used: MIMIC-III, MIMIC-IV, and eICU-CRD.

#### MIMIC-III

MIMIC-III (Medical Information Mart for Intensive Care III) is a comprehensive and publicly accessible database that contains de-identified health data associated with over 40,000 thousand patients who stayed in critical care units of the Boston-based Beth Israel Deaconess Medical Center (BIDMC) between 2001 and 2012. It is maintained by the Laboratory for Computational Physiology (LCP) at MIT and is shared through the PhysioNet platform. The database includes information such as demographics, vital sign measurements, laboratory test results, medications, and more. Since its release in 2016, it has served as a valuable resource for a wide range of research studies in healthcare, including those focused on critical care and machine learning applications in medicine. ^6^

#### MIMIC-IV

MIMIC-IV builds on the foundation laid by MIMIC-III, extending the dataset to include patients admitted to the ICU from 2008 to 2019. Unique features of MIMIC-IV include clinical progress notes and physiological data collected from bedside monitors. Approximately 70,000 de-identified medical records are archived in the MIMIC-IV database. ^7^

As there is potential overlap of patients between 2008–2012 across both MIMIC versions, where the same patient may have distinct but not linkable identifiers, users of our dataset may consider dropping MIMIC-III encounters entirely or restricting their analysis to those corresponding to 2013-2019.

##### eICU-CRD

The eICU-CRD database is a publicly available multi-center database sourced from the Philips Healthcare eICU Telehealth Program. It contains information about over 200,000 admissions from 208 hospitals or ICUs monitored by eICU programs across the United States, between 2014 and 2015. The eICU-CRD patients are distinct from MIMIC-III and MIMIC-IV subjects, alleviating any concerns regarding the potential overlap of their underlying populations. ^5^

### Software

For data extraction, BigQuery through Google Colaboratory (Python 3.10) was used.

### Publicly-available derived views

The derived tables available on the MIMIC-Code (*icustay_detail*, *vitalsign*, *complete_blood_count*, *coagulation*, *chemistry*, *bg*, *enzyme*, and *sofa*) and eICU-Code (*pivoted_vital* and *pivoted_lab*) repositories, made available on the MIT-LCP GitHub repository, were used. ^9^

### Inclusion and exclusion criteria

All patients admitted to a hospital or ICU captured by one of the aforementioned databases who had valid ABG and pulse oximetry data were included. Two versions of the dataset were obtained: an extended dataset created primarily for validation purposes and a preprocessed, validated dataset shared in the present study:

1. Extended dataset (mainly for technical validation purposes):

– (SaO_2_, SpO_2_) pairs are captured within 90 minutes
– No range for the oxygen saturation is set
– All pairs per hospital admission are considered
2. Preprocessed dataset (the shared version):

– (SaO2, SpO_2_) pairs are captured within 5 minutes ^10,11^
– A range of 70-100% is set
– Only the first pair per hospital admission is considered

### SaO_2_ – SpO_2_ matching

We require each pulse oximetry reading (SpO_2_) to precede the ABG measurement (SaO_2_). Missing ABG data is not allowed. For the extended dataset, the window is [-90, 0] minutes; for the preprocessed dataset, [-5, 0] minutes. Figure 1 depicts the rationale of the final dataset.

**Figure 1.**
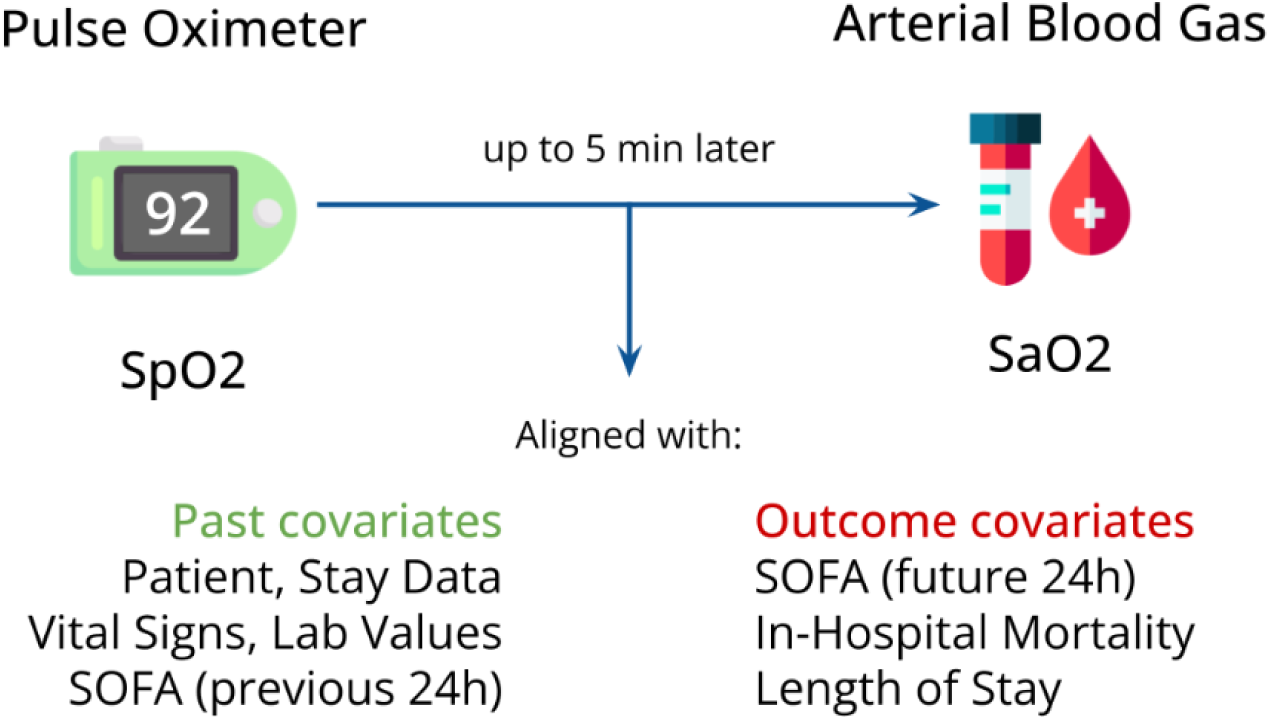
Rationale and variables included in the dataset.

### Time alignment and curation across different databases

To facilitate modifications, ensure that definitions remain consistent across databases, and promote subsequent code reuse by other teams, we followed these steps to align each (SaO_2_, SpO_2_) pair with time-varying covariates:

1. Create pivoted views of the lab measurements, vital signs, and hourly SOFA scores (either publicly-available on BigQuery, or generated by our team);
2. List the variables to be pulled, each with the following fields:

a. Variable type (for the prefix)
b. Original name (for the pull)
c. New name (harmonized across databases)
d. Time window for the value to be considered (variable-specific)
e. Source table (the *id* of the pivoted view)
f. Used foreign key (that links the (SaO_2_, SpO_2_) pair with the source table)
g. Name of the timestamp variable (specific to the source table)
3. Parse the new table (saved as an editable *Google Spreadsheet*) through *Pandas,* on Google Colab
4. Create the complete SQL query automatically, feeding it with all listed variables, previously aligned with the (SaO_2_, SpO_2_) pairs through separate subqueries
5. Query the databases through the *Python BigQuery API*

The above-described stepwise process is summarized in Figure 2. By setting relevant time windows for each variable, we ensure to extract relevant data only. For example, a temperature reading will only be aligned with a (SaO_2_, SpO_2_) pair if registered up to 8 hours before the SaO_2_value was measured. Missingness is kept as is to give users the flexibility to adopt their own imputation strategy, if needed.

**Figure 2.**
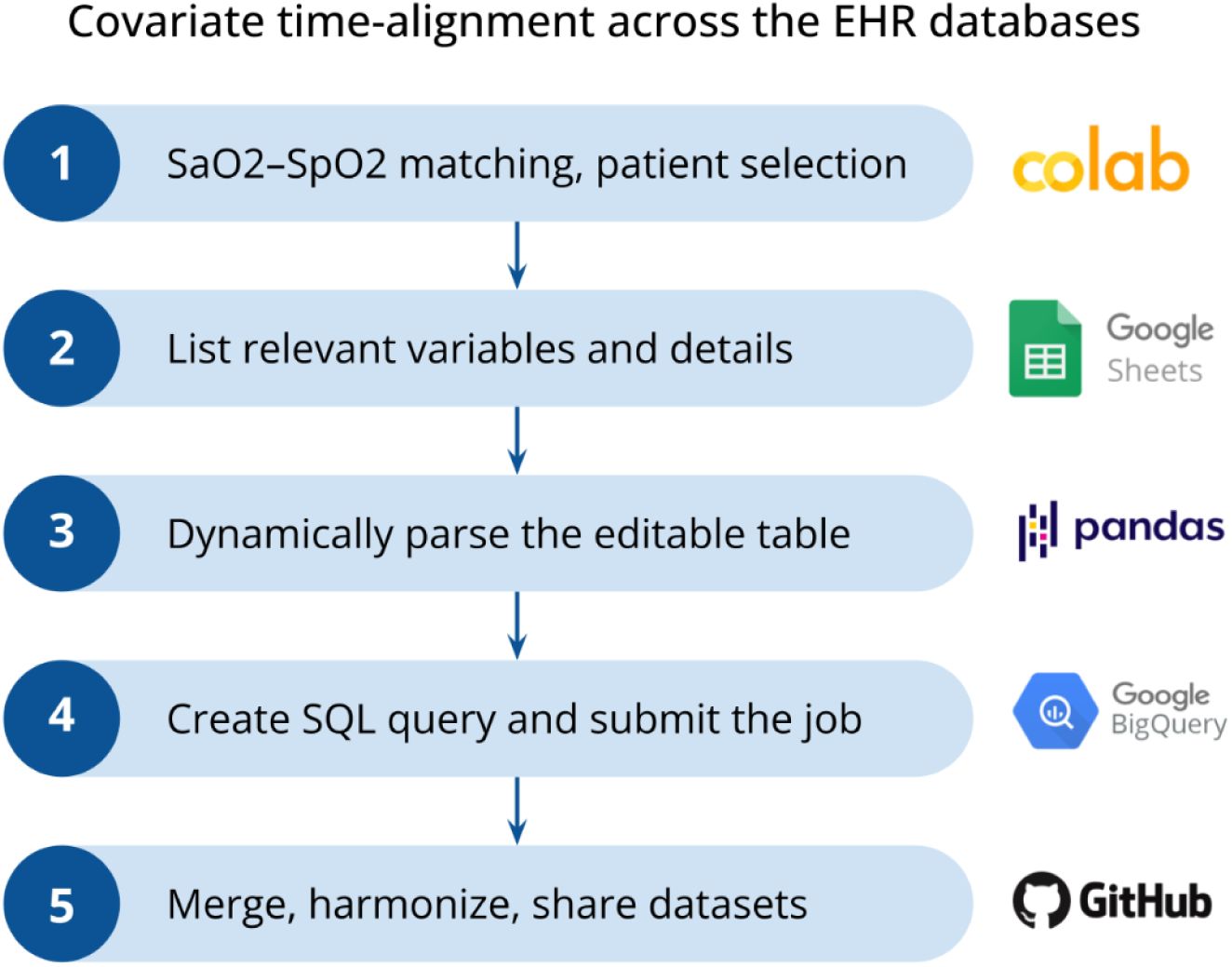
Pipeline created to curate and merge the datasets.

**Figure 3a.**
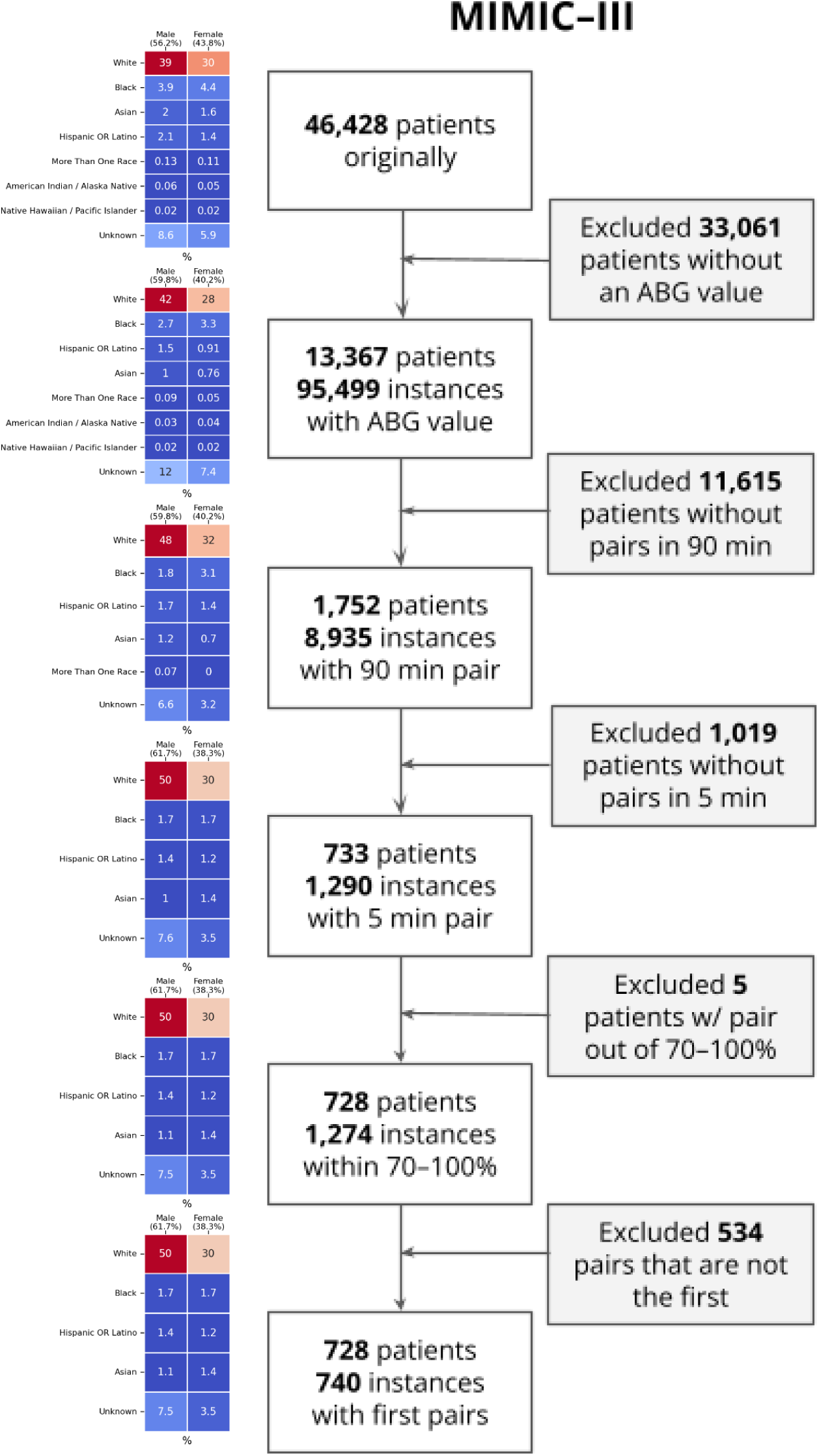
Flow diagram for MIMIC-III depicting cohort selection.

**Figure 3b.**
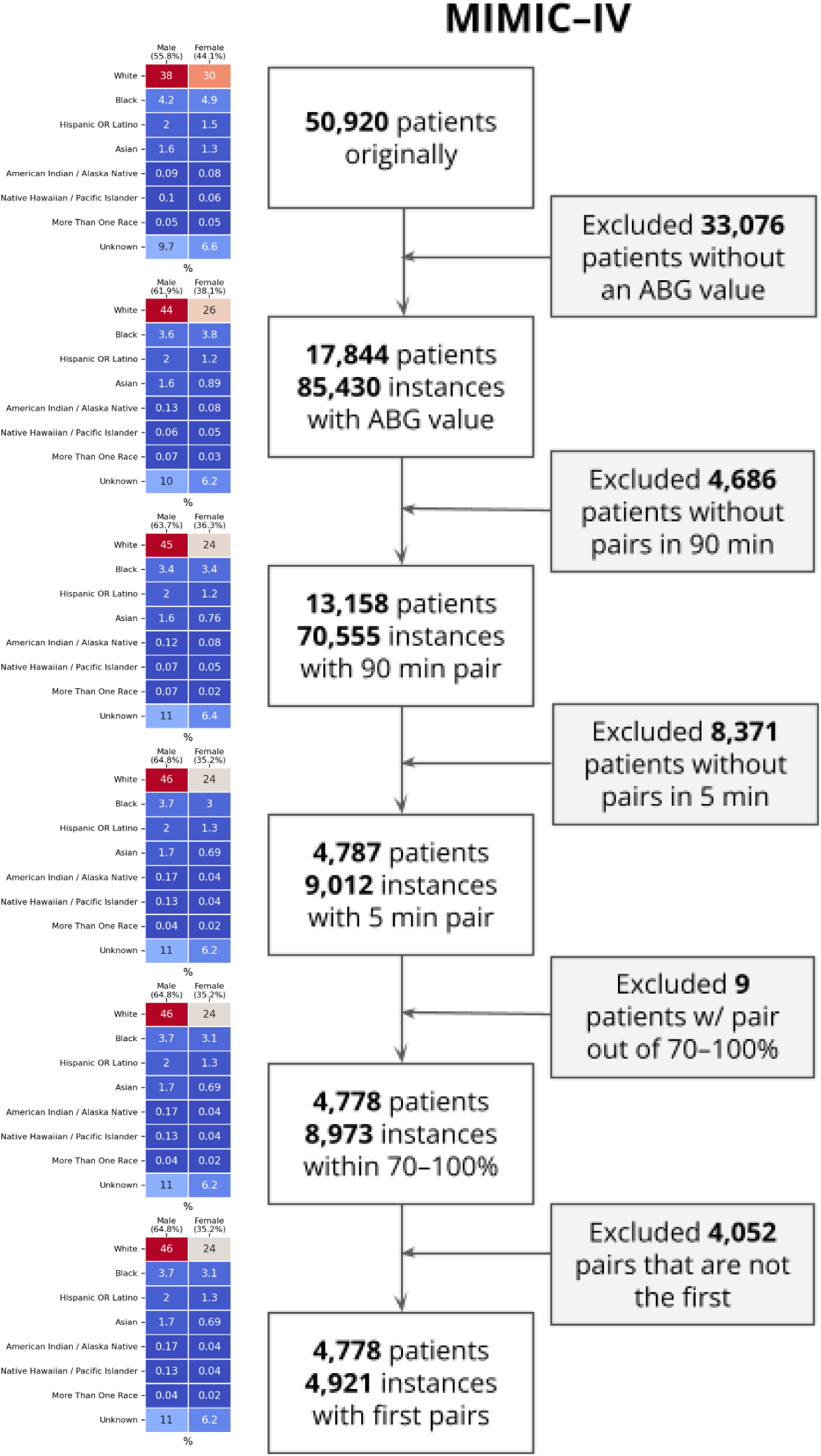
Flow diagram for MIMIC-IV depicting cohort selection.

**Figure 3c.**
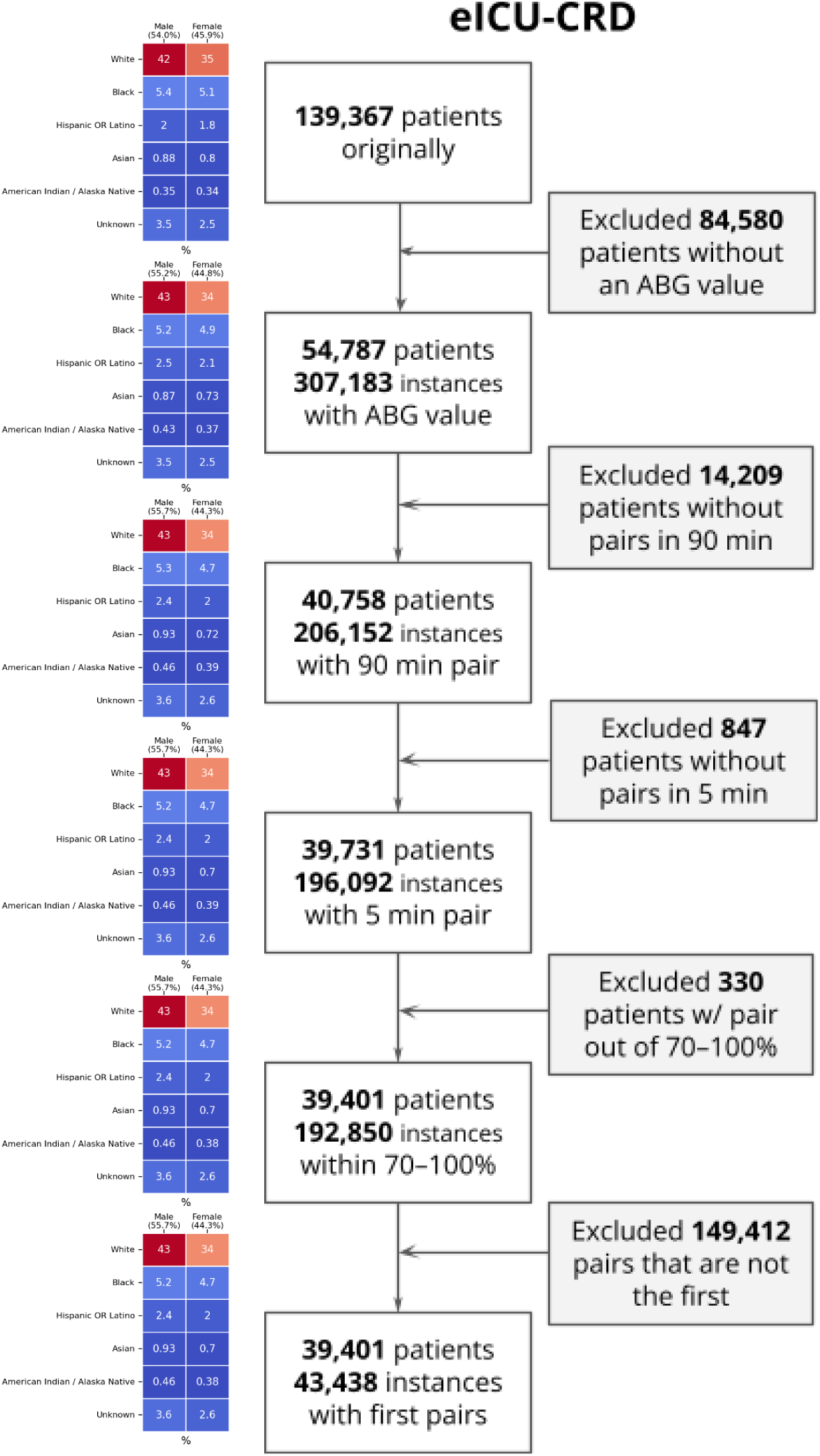
Flow diagram for eICU-CRD depicting cohort selection.

**Figure 4.**
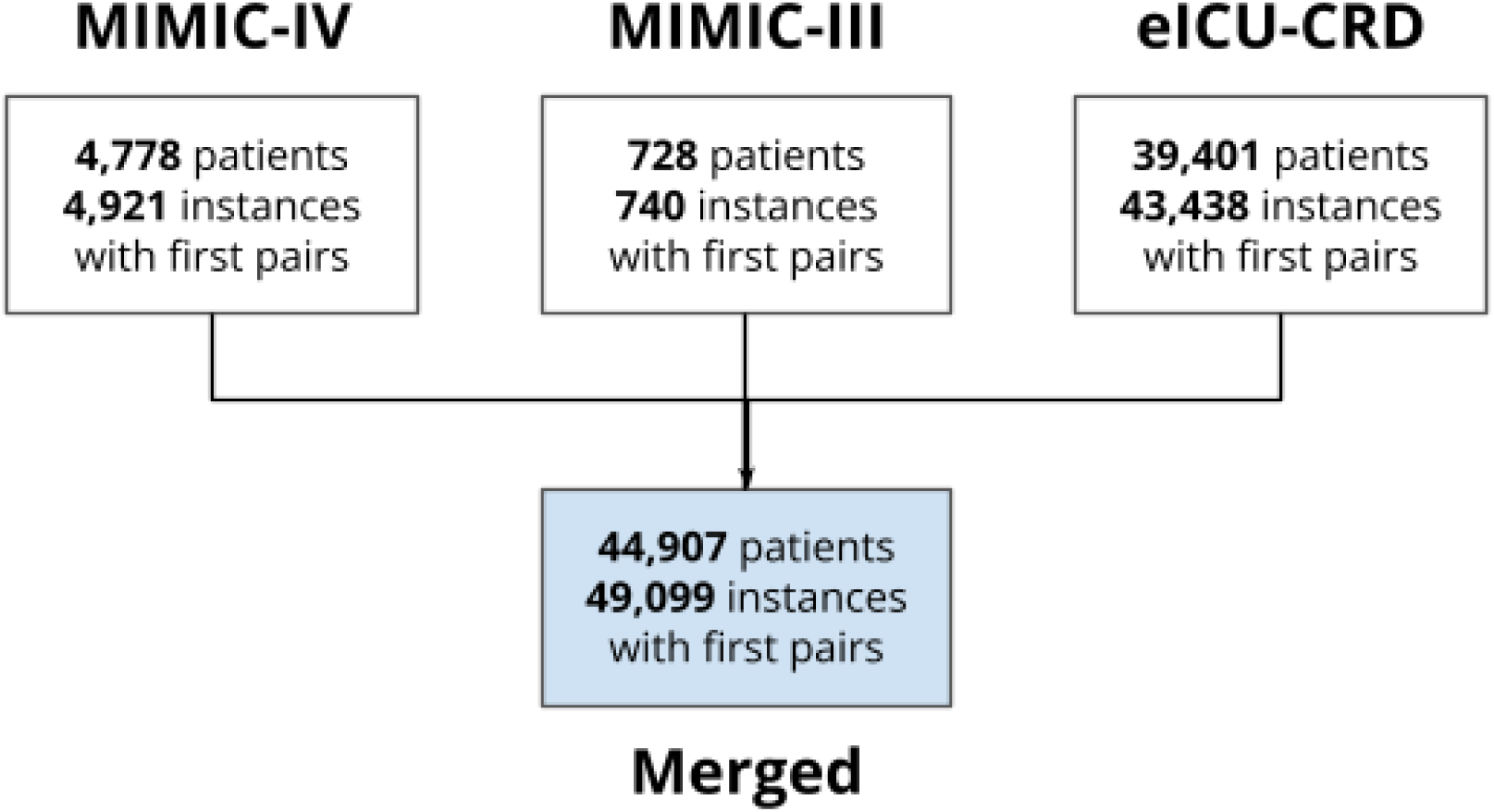
Flow diagram for the merged dataset.

### Harmonization of concepts among databases

Data from different tables were harmonized into the same format across all databases. To minimize missingness, only variables that are available in all three databases were included. Patient-level variables (*identifiers*; *demographics; admission characteristics* and *patient outcomes*) were unified as shown in the Supplemental Table 1. Time-varying variables (*vital signs*; *laboratory test values*; *hourly SOFA scores*) were pulled and harmonized as shown in Table 1. Each variable maps to an *itemid* (respectively, *label*) in the MIMIC (respectively, eICU-CRD) databases.

**Table 1.**
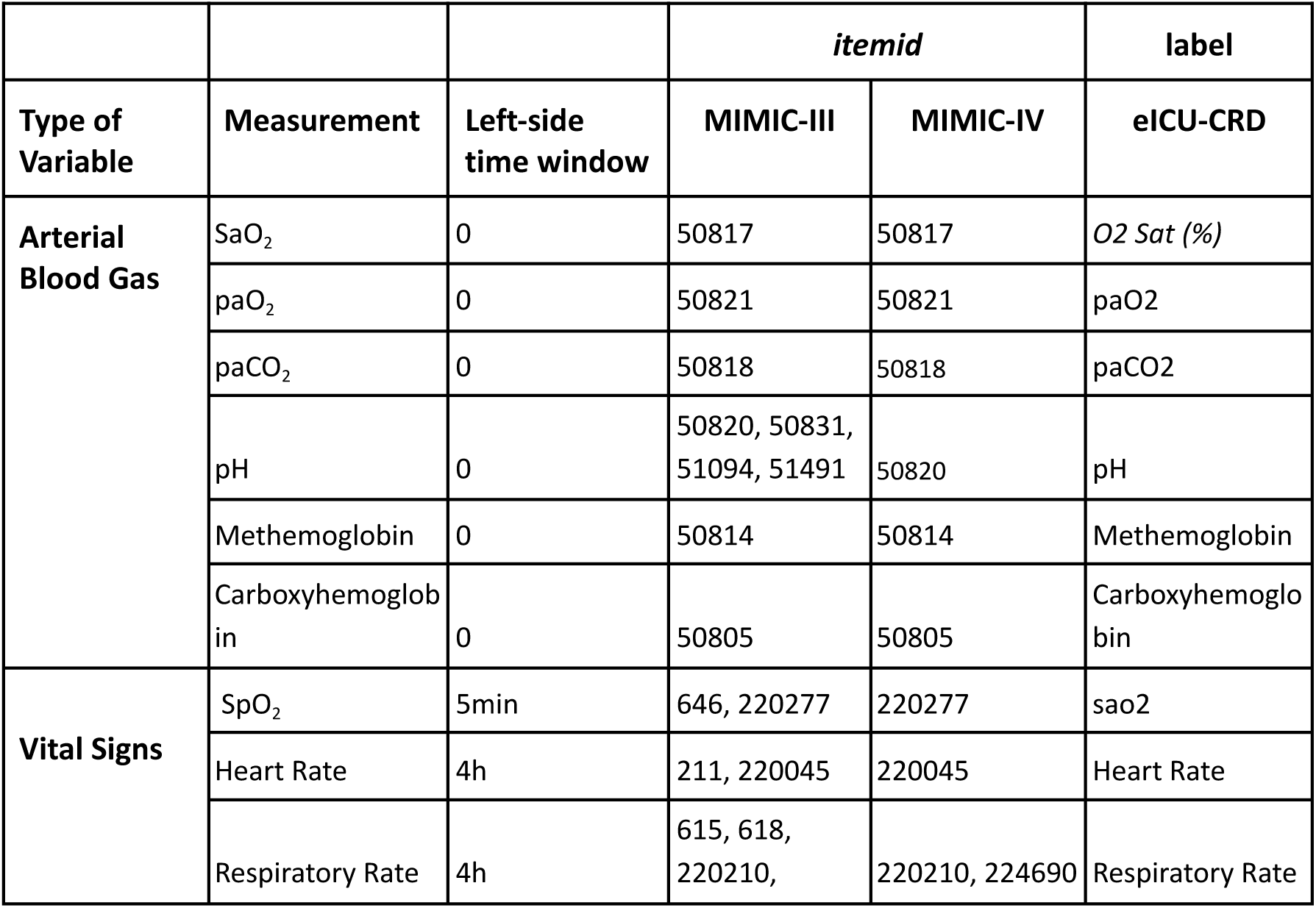
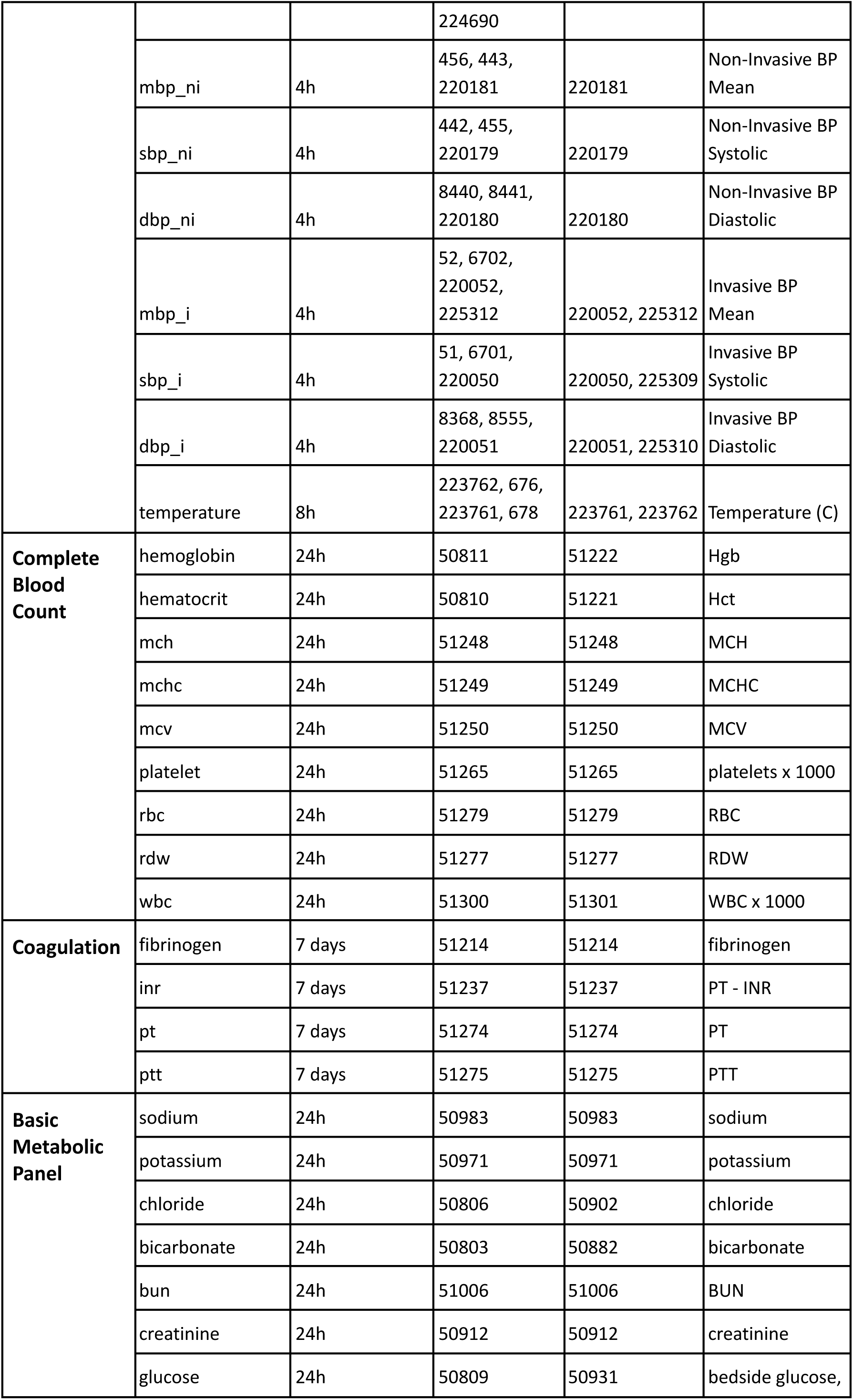
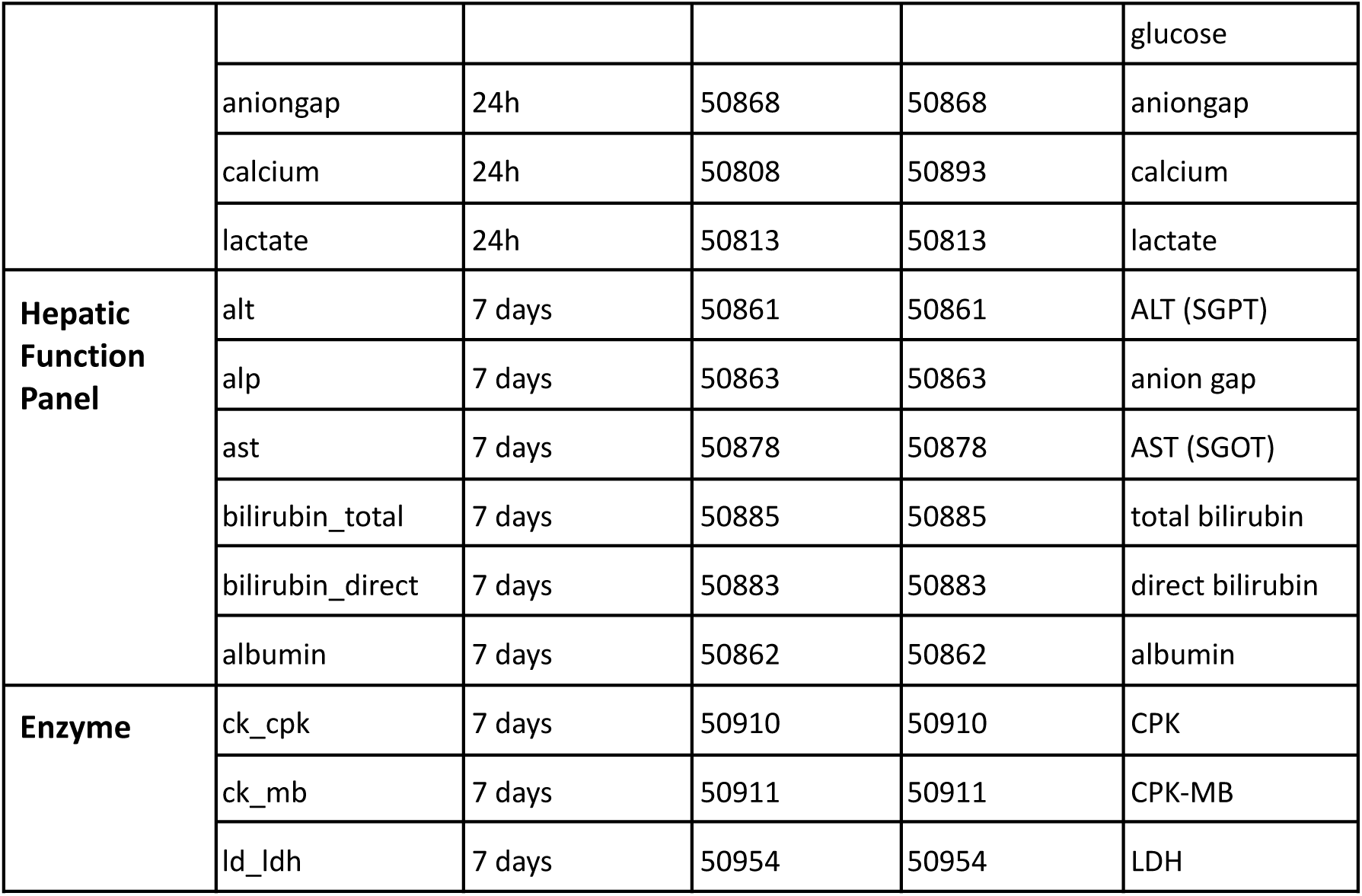
Item IDs related to SaO_2_ and other laboratory test values, stratified by source database.

|  |  |  | itemid |  | label |
| --- | --- | --- | --- | --- | --- |
| Type of Variable | Measurement | Left-side time window | MIMIC-III | MIMIC-IV | eICU-CRD |
| Arterial Blood Gas | SaO<sub>2</sub> | 0 | 50817 | 50817 | O2 Sat (%) |
|  | paO<sub>2</sub> | 0 | 50821 | 50821 | paO2 |
|  | paCO<sub>2</sub> | 0 | 50818 | 50818 | paCO2 |
|  | pH | 0 | 50820, 50831, 51094, 51491 | 50820 | pH |
|  | Methemoglobin | 0 | 50814 | 50814 | Methemoglobin |
|  | Carboxyhemoglobin | 0 | 50805 | 50805 | Carboxyhemoglobin |
| Vital Signs | SpO<sub>2</sub> | 5min | 646, 220277 | 220277 | sao2 |
|  | Heart Rate | 4h | 211, 220045 | 220045 | Heart Rate |
|  | Respiratory Rate | 4h | 615, 618, 220210, | 220210, 224690 | Respiratory Rate |
|  |  |  | 224690 |  |  |
|  | mbp_ni | 4h | 456, 443, 220181 | 220181 | Non-Invasive BP Mean |
|  | sbp_ni | 4h | 442, 455, 220179 | 220179 | Non-Invasive BP Systolic |
|  | dbp_ni | 4h | 8440, 8441, 220180 | 220180 | Non-Invasive BP Diastolic |
|  | mbp_i | 4h | 52, 6702, 220052, 225312 | 220052, 225312 | Invasive BP Mean |
|  | sbp_i | 4h | 51, 6701, 220050 | 220050, 225309 | Invasive BP Systolic |
|  | dbp_i | 4h | 8368, 8555, 220051 | 220051, 225310 | Invasive BP Diastolic |
|  | temperature | 8h | 223762, 676, 223761, 678 | 223761, 223762 | Temperature (C) |
| **Complete Blood Count** | hemoglobin | 24h | 50811 | 51222 | Hgb |
|  | hematocrit | 24h | 50810 | 51221 | Hct |
|  | mch | 24h | 51248 | 51248 | MCH |
|  | mchc | 24h | 51249 | 51249 | MCHC |
|  | mcv | 24h | 51250 | 51250 | MCV |
|  | platelet | 24h | 51265 | 51265 | platelets x 1000 |
|  | rbc | 24h | 51279 | 51279 | RBC |
|  | rdw | 24h | 51277 | 51277 | RDW |
|  | wbc | 24h | 51300 | 51301 | WBC x 1000 |
| **Coagulation** | fibrinogen | 7 days | 51214 | 51214 | fibrinogen |
|  | inr | 7 days | 51237 | 51237 | PT - INR |
|  | pt | 7 days | 51274 | 51274 | PT |
|  | ptt | 7 days | 51275 | 51275 | PTT |
| **Basic Metabolic Panel** | sodium | 24h | 50983 | 50983 | sodium |
|  | potassium | 24h | 50971 | 50971 | potassium |
|  | chloride | 24h | 50806 | 50902 | chloride |
|  | bicarbonate | 24h | 50803 | 50882 | bicarbonate |
|  | bun | 24h | 51006 | 51006 | BUN |
|  | creatinine | 24h | 50912 | 50912 | creatinine |
|  | glucose | 24h | 50809 | 50931 | bedside glucose, glucose |
|  | aniongap | 24h | 50868 | 50868 | aniongap |
|  | calcium | 24h | 50808 | 50893 | calcium |
|  | lactate | 24h | 50813 | 50813 | lactate |
| **Hepatic Function Panel** | alt | 7 days | 50861 | 50861 | ALT (SGPT) |
|  | alp | 7 days | 50863 | 50863 | anion gap |
|  | ast | 7 days | 50878 | 50878 | AST (SGOT) |
|  | bilirubin_total | 7 days | 50885 | 50885 | total bilirubin |
|  | bilirubin_direct | 7 days | 50883 | 50883 | direct bilirubin |
|  | albumin | 7 days | 50862 | 50862 | albumin |
| **Enzyme** | ck_cpk | 7 days | 50910 | 50910 | CPK |
|  | ck_mb | 7 days | 50911 | 50911 | CPK-MB |
|  | ld_ldh | 7 days | 50954 | 50954 | LDH |

Since eICU-CRD is designed around time offsets (e.g., minutes from ICU admission, minutes from hospital admission, etc.), all dates were converted from offsets by a reference date of January 1^st^, 2014 (since eICU-CRD encompasses 2014-2015 data).

For race and ethnicity, the NIH Policy on Reporting Race and Ethnicity was used as the reference.^12^ The eight unified categories were: “*American Indian / Alaska Native”*, *“Asian”*, *“Black or African American”*, *“Hispanic OR Latino”*, *“More Than One Race”*, *“Native Hawaiian / Pacific Islander”*, *“White”*, *and “Unknown”*. We mapped the original race and ethnicity labels present in the databases we studied to these categories. The exact mappings are further depicted in Supplemental Tables 2a, 2b, and 2c. Based upon how data was captured, none of the databases are able to distinguish between race (e.g., Asian, Black, White) and ethnicity (e.g., “Hispanic OR Latino” vs “Not Hispanic or Latino”). We denote “Hispanic OR Latino” as “Hispanic”, a coded value. As such, a patient could be addressed as either any race (but non-Hispanic ethnicity) or Hispanic ethnicity (of any race). This limitation remains present in BOLD.

### Data types

Several additional variables can help augment analyses and characterize patients receiving a temporally proximate (SaO_2_, SpO_2_) pair. These are further described below.

All adjunctive (e.g., vital signs, laboratory test values, etc.) data are referenced from the time of the ABG. For the purpose of this manuscript, a time delta (*delta_* prefix) refers to the time difference between the time of the most recently recorded covariate of interest and that of the ABG measurement. Each time-varying covariate is accompanied by a time delta. The ABG measurement time is set as the reference; any covariate measurement or reading must occur before this reference, unless otherwise noted.

#### Identifiers

Each encounter has three identifiers, at different levels: patient, hospital, and ICU admission. The original identifiers are kept to allow linking the data with the original databases and eventually pull other variables of interest. However, to avoid overlap among the databases, we created new, unique identifiers for our preprocessed dataset; they reflect each of these three identifiers. In addition, each encounter has an identifier to reflect the source database.

Among the three considered databases, only eICU-CRD has hospital identifiers, since the MIMIC databases come from one single hospital, i.e., BIDMC. As a result, MIMIC data was assigned a hospital index of *9999*, which is outside the range of eICU-CRD hospital indices. Other hospital-related variables (number of beds, US region, and teaching status) were harmonized accordingly.

#### Demographics

Demographics, such as age at admission, sex, race and ethnicity, were extracted from the demographics tables of each database. Age at admission age was unified, with values between 18-89 kept intact and values of 90 and above taken equal to 90.

#### Admission characteristics and patient outcomes

Comorbidities are calculated by van Walraven Elixhauser score ^13^ (MIMIC-III) and Charlson Comorbidity Index ^14^ (MIMIC-IV, eICU-CRD). BMI was computed with the weight and height on admission, for each database.

Hospital-level (e.g., hospital size) and patient-specific admission characteristics (e.g., admission time) as well as patient-specific outcomes (e.g., in-hospital mortality) were recorded for each encounter.

#### Vital signs

Vital sign data were merged in accordance with Table 1. Temperature, blood pressure (both non-invasive and invasive), heart rate, respiratory rate, and SpO_2_ were extracted. These data were obtained from the *chartevents* and *nursecharting* tables of the original MIMIC and eICU databases, respectively. The prefix “*vitals_*” is used for each variable of this type, except for SpO_2_.

#### Laboratory test values

Common laboratory test values were merged within variable-specific time windows as noted. Measurements of the following categories were pulled: ABG (no prefix), complete blood count (“*cbc_”* prefix); coagulation (“*coag_”* prefix); basic metabolic panel (“*bmp_”* prefix); hepatic function panel (“*hfp_”* prefix); and other enzymes (“*other_”* prefix). In the MIMIC databases, all laboratory test data were collected from the original *labevents* table; in eICU-CRD, data were collected from the *labs* table.

#### Hourly SOFA scores

To characterize organ dysfunction and severity of illness, sequential organ failure assessment (SOFA) scores were used.^15^ SOFA scores for each dataset were calculated hourly. SOFA scores were extracted in the hour prior to the ABG to ensure that the latter has no impact on characterizing underlying organ dysfunction and thus avoid reverse causation (*“sofa_past_”* prefix). To quantify the impact of hypoxemia on organ dysfunction, subsequent SOFA scores were also extracted 24 hours after the ABG (*“sofa_future_”* prefix).

In the MIMIC databases, the publicly available derived table with hourly SOFA scores were used. In the eICU-CRD, we used an auxiliary query created by our team.

### Data storage

The preprocessed dataset, meeting the defined inclusion / exclusion criteria, is stored on PhysioNet as a single comma separated value (CSV) file.

### Descriptive analytics and technical validation

We now present the methodology followed to support the criteria we set to select patients and clean the data.

Flow diagrams depicting the application of inclusion and exclusion criteria to select our cohort were created and analyzed. At each exclusion step, we analyzed the composition of the patients who are dropped in terms of demographics.

We created descriptive tables highlighting patients’ characteristics across source databases; race and ethnicity; and hidden hypoxemia – when SpO_2_ ≥ 88% but SaO_2_< 88%, as defined by Wong et al. ^2^ The *tableone* package was used. ^16^

We employed Modified Bland-Altman plots, based on the methodology proposed by Wong et al. ^2^, to evaluate the agreement between SaO_2_and SpO_2_ measurements. We assessed the calibration performance across two different time window sizes — 5 and 30 minutes — to justify our final selection of a 5-minute window. Moreover, we conducted separate analyses across racial and ethnic groups to highlight disparities in calibration accuracy.

Oxyhemoglobin dissociation curves ^17^ are also reported as a referential integrity of our ABG data. To verify the existence of left and right shifts, we plotted, in different colors, the pairs with pH in the 90th and 10th percentiles, respectively.

The root mean squared error (RMSE) of each (SaO_2_, SpO_2_) pair was computed across different window limits, from 0 to 90, and stratified by race and ethnicity (for simplicity, considered groups were White, Black, Hispanic OR Latino, and Asian). RMSE was computed using equation (1) for each pair, aggregated with a mean, and then 95% confidence intervals were computed assuming normal distributions.

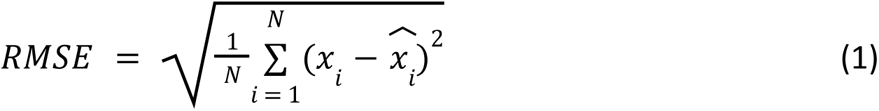

Finally, the *missingno* package ^18^ was used to assess the completeness of the data, reported as a bar plot.

## Data Records

### Description of fields

#### Demographics

***subject_id*:** Describes a unique subject. This is unique per component dataset and mapped directly to the equivalent term in each. A subject may have multiple admissions, denoted by hospital_admission_id. Same as in the original database.

***hospital_admission_id*:** Describes a unique hospital admission. This is unique per component dataset and mapped directly to the equivalent term in each. Same as in the original database.

***icustay_id*:** Describes a unique ICU admission. This is unique per component dataset and mapped directly to the equivalent term in each. Each hospital admission may have multiple ICU stays. Same as in the original database.

***unique_subject_id*:** Describes a unique subject. Guarantees that no subjects coming from different databases have the same identifier.

***unique_hospital_admission_id*:** Describes a unique hospital admission. Guarantees that no subjects coming from different databases have the same identifier.

***unique_icustay_id*:** Describes a unique ICU admission. Guarantees that no subjects coming from different databases have the same identifier.

***source_db*:** Labeled as *mimic_iii*, *mimic_iv*, *eicu* to distinguish between component datasets.

***race_ethnicity*:** Harmonized race and ethnicity.

#### Hospital characteristics

***hospitalid*:** Unique hospital ID. Beth Israel Deaconess (MIMIC-III, −IV) was denoted as 9999.

***numbedscategory*:** Hospital size, in numbers of beds. This was recorded as “*< 100*”; “*100-250*”; “*250-500*”; “*≥ 500*” beds for eICU-CRD. For MIMIC-III and −IV (BIDMC), we fixed the value at “*≥ 500” beds*.

***teachingstatus*:** This field marks whether a hospital was identified as a teaching hospital, where a value of 1 implies that it was a teaching hospital. BIDMC has teaching status = 1.

***region*:** Maps to the US census region distribution per eICU. It is either *Midwest, Northeast*, *South*, or *West.* MIMIC (BIDMC) was set as *Northeast*.

***admission_age*:** Harmonized age on admission. eICU admission age mapped to admission_age. MIMIC-III admission age mapped to admission age. MIMIC-IV admission_age mapped to admission_age.

***sex_female*:** Assigned a value of 1 if the patient is of female sex. ***weight_admission*:** Weight on admission (in kilograms). ***height_admission*:** Height on admission (in centimeters).

***BMI_admission*:** Calculated BMI on admission, based on weight_admission and height_admission. If either weight_admission or height_admission is missing, BMI_admission is missing.

#### Admission characteristics

***datetime_hospital_admit, datetime_hospital_discharge, datetime_icu_admit, datetime_icu_discharge*:** Date and time of hospital and ICU admission (_admit) and discharge (_discharge).

***los_hospital, los_ICU*:** Length of stay for hospital (_hospital) and ICU (_ICU) in days.

***in_hospital_mortality*:** This variable is true if the patient died during the hospital admission, regardless if the patient died during the ICU admission or not.

***comorbidity_score_name, comorbidity_score_value*:** Comorbidity score (either Elixhauser or Charlson), along with score.

#### ABG data

***SaO2_timestamp*:** Date and time of ABG test.

***pH***: pH value.

***pCO2*:** Partial pressure of CO2 (mmHg).

***pO2***: Partial pressure of O2 (mmHg).

***SaO2*:** Arterial oxygen saturation (%).

***Carboxyhemoglobin*:** Percentage of hemoglobin bound to CO (%).

***Methemoglobin*:** Percentage of methemoglobin (%).

#### Vitals data

***SpO2*:** Pulse oximetry saturation (%).

***SpO2_timestamp*:** Date and time of SpO_2_ recording.

***vitals_heart_rate*:** Heart rate.

***vitals_resp_rate*:** Respiratory rate.

***vitals_mbp_ni, vitals_sbp_ni, vitals_dbp_ni:*** Mean arterial pressure (MAP), Systolic pressure (SBP), Diastolic pressure (SBP) calculated from noninvasive (cuff) blood pressure.

***vitals_mbp_i, vitals_sbp_i, vitals_dbp_i:*** Mean arterial pressure (MAP), Systolic pressure (SBP), and Diastolic pressure (DBP) calculated from invasive (arterial line) blood pressure.

***vitals_tempc:*** Temperature, from any body measuring site, in Celsius (°C).

#### Labs

##### Complete Blood Count (CBC)

***cbc_wbc*:** White blood cell. (10^9^/L)

***cbc_hemoglobin*:** Measured hemoglobin (g/L).

***cbc_hematocrit*:** Measured hematocrit. (%)

***cbc_platelet*:** Measured platelet count.(10^9^/L)

***cbc_mch*:** Measured mean corpuscular hemoglobin. (pg)

***cbc_mchc*:** Measured mean corpuscular hemoglobin concentration. (g/L)

***cbc_mcv*:** Measured mean corpuscular volume.(fL) ***cbc_rbc*:** Measured red blood cells (RBC). (10^12^/L) ***cbc_rdw*:** Measured RBC distribution width. (%)

##### Coagulation labs

***coag_fibrinogen*:** Measured fibrinogen.

***coag_pt*:** Measured prothrombin time. (s)

***coag_inr*:** Measured international normalized ratio.

***coag_ptt*:** Measured partial thromboplastin time. (s)

##### Basic Metabolic Panel (BMP)

***bmp_sodium*:** Measured sodium levels. (mmol/L)

***bmp_potassium*:** Measured potassium levels. (mmol/L)

***bmp_chloride*:** Measured chloride levels. (mmol/L)

***bmp_bicarbonate*:** Measured bicarbonate levels. (mg/dL)

***bmp_bun*:** Measured blood urea nitrogen levels. (mg/dL)

***bmp_creatinine*:** Measured creatinine levels. (mg/dL)

***bmp_glucose*:** Measured glucose levels. (mg/dL)

***bmp_aniongap*:** Measured anion gap. (mmol/L)

***bmp_calcium*:** Measured calcium levels. (mg/dL)

***bmp_lactate*:** Measured lactate levels. (mmol/L)

##### Hepatic function panel (HFP)

***hfp_alt*:** Measured alanine aminotransferase (ALT) levels. (U/L)

***hfp_alp*:** Measured alkaline phosphatase (ALP) levels. (U/L)

***hfp_ast*:** Measured aspartate aminotransferase (AST) levels. (U/L)

***hfp_bilirubin_total*:** Measured total bilirubin levels. (mg/dL)

***hfp_bilirubin_direct*:** Measured direct bilirubin levels. (mg/dL)

***hfp_albumin*:** Measured albumin levels. (g/dL)

##### Other labs (enzyme)

***others_ck_cpk*:** Measured creatine kinase (CK) levels, also known as creatine phosphokinase (CPK). (U/L)

***others_ck_mb*:** Measured creatine kinase MB (CK-MB) levels. (U/L)

***others_ld_ldh*:** Measured lactate dehydrogenase (LDH) levels. (U/L)

#### SOFA scores

***sofa_past_overall_24hr*:** SOFA score, calculated from component values below, measured in the hour window prior to the ABG.

***sofa_past_coagulation_24hr, sofa_past_liver_24hr, sofa_past_cardiovascular_24hr, sofa_past_cns_24hr, sofa_past_renal_24hr*:** SOFA score components, with highest value for each component in the past 24 hours. The hour window just prior to the hour window containing the ABG is recorded here to characterize baseline patient status.

***sofa_future_overall_24hr*:** SOFA score, calculated from component values below, measured 24 hours after the ABG window.

***sofa_future_coagulation_24hr, sofa_future_liver_24hr, sofa_future_cardiovascular_24hr, sofa_future_cns_24hr, sofa_future_renal_24hr*:** SOFA score components, with highest value for each component in the 24 hours after the ABG to characterize the 24 hour impact of discrepancies.

### Technical Validation

In the extended dataset, at the loose cut-off of 90 minutes, we obtained on average 5 pairs for *Asian* and *Hispanic OR Latino* patients, 4.7 pairs for *Black* and *American Indian / Alaska Native* patients, and 4.5 pairs for *White* patients (see Table 2 with the average number of pairs per race and ethnicity).

**Table 2.** Average number of pairs per race and ethnicity, extended dataset.

| Race and Ethnicity | Average No. Pairs (Standard Deviation) ↓ | N |
| --- | --- | --- |
| White | 4.47 (7.74) | 43,925 |
| Black | 4.65 (8.23) | 5,467 |
| American Indian / Alaska Native | 4.71 (8.49) | 397 |
| Hispanic OR Latino | 5.09 (8.65) | 2,436 |
| Asian | 5.12 (10.88) | 1,061 |
| Unknown | 4.96 (9.47) | 5,030 |

To ensure that the distributions of SaO_2_ and PaO_2_ values obtained in the three considered EHR were concordant with the literature, we plotted an oxyhemoglobin dissociation curve (Figure 5). We did not observe substantial deviation from the known dissociation curve. Specifically, the pH-associated left and right shifts were verified, ensuring that the data curation process in the original databases was not flawed.

**Figure 5.**
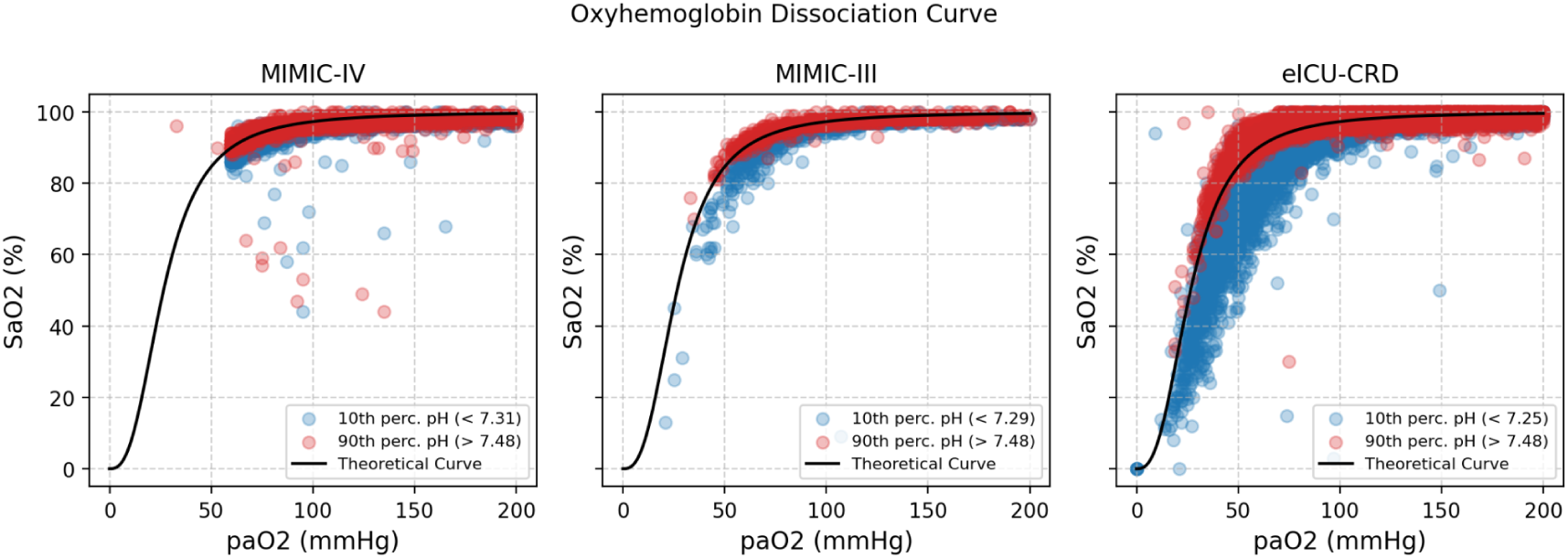
Oxyhemoglobin dissociation curve, per database, with the pH shift highlighted, on the extended dataset.

We examined the pairs of the extended dataset to study the agreement of SpO_2_ measurements by pulse oximeter with the SaO_2_ measurements by ABG. We assessed various lengths of the eligible time window (*delta_SpO2*) to pair readings. The modified Bland-Altman plots presented in Figure 6 revealed no significant differences between the two readings over time windows of length 5 and 30 minutes, respectively. The patterns remained the same irrespective of the database, patient race, and patient ethnicity. However, increasing *delta_SpO2* tolerances yielded a sharp increase in the RMSE for Asian, Black, and Hispanic patients (Figure 7); this change was most pronounced for a time delta of 60 minutes or more, most likely due to lower sample sizes among patients from minority groups than among White patients. Although increasing the time delta mechanistically resulted in an incremental increase in the number of eligible paired samples, we selected 5 minutes as the optimal cut-off. Indeed, it coincided with a marked increase in paired samples, while still yielding a relatively small RMSE.

**Figure 6.**
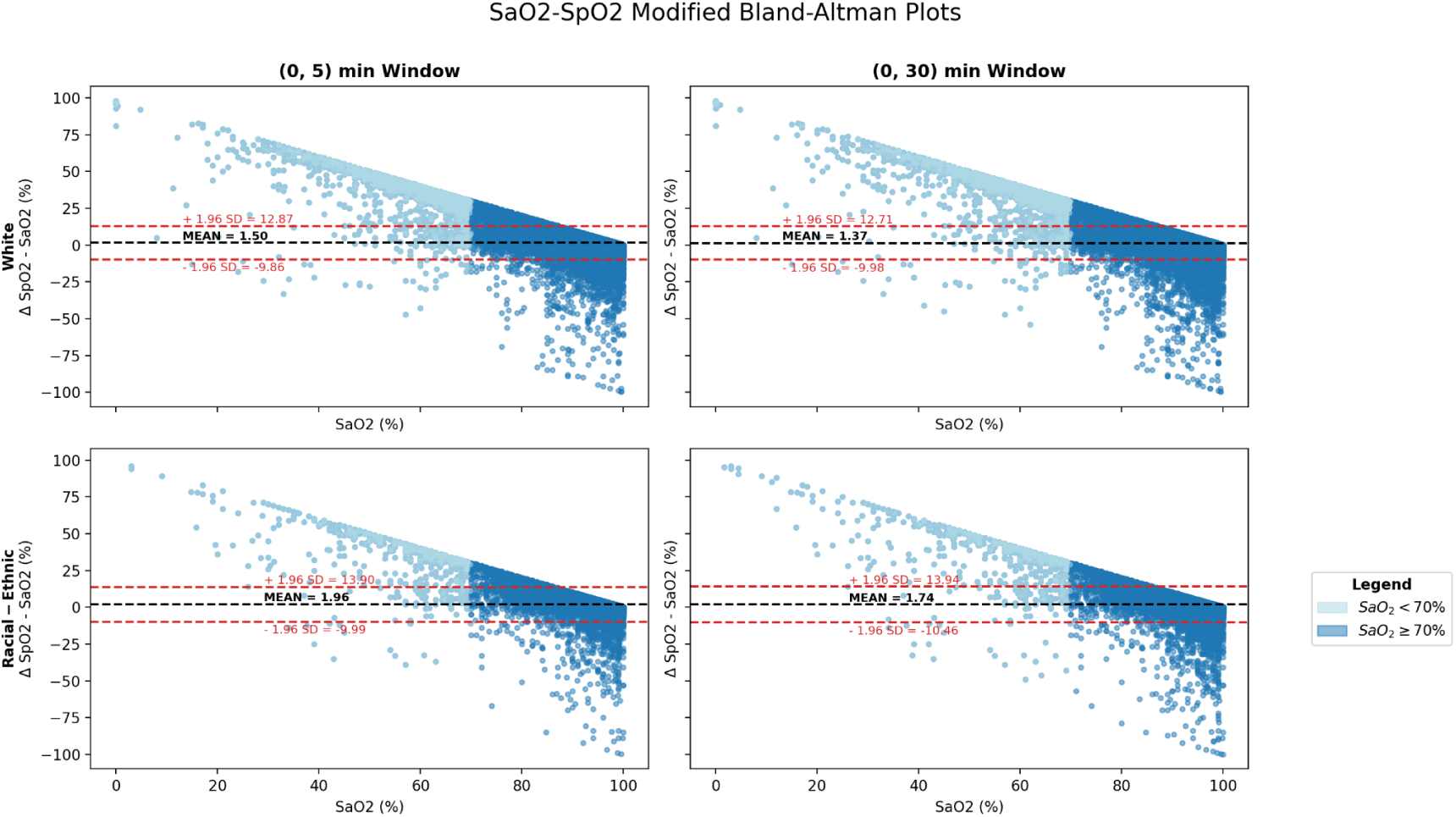
Modified Bland-Altman plots, across race and ethnicity (White compared with racial and ethnic group), and across 2 time windows, on the extended dataset.

**Figure 7.**
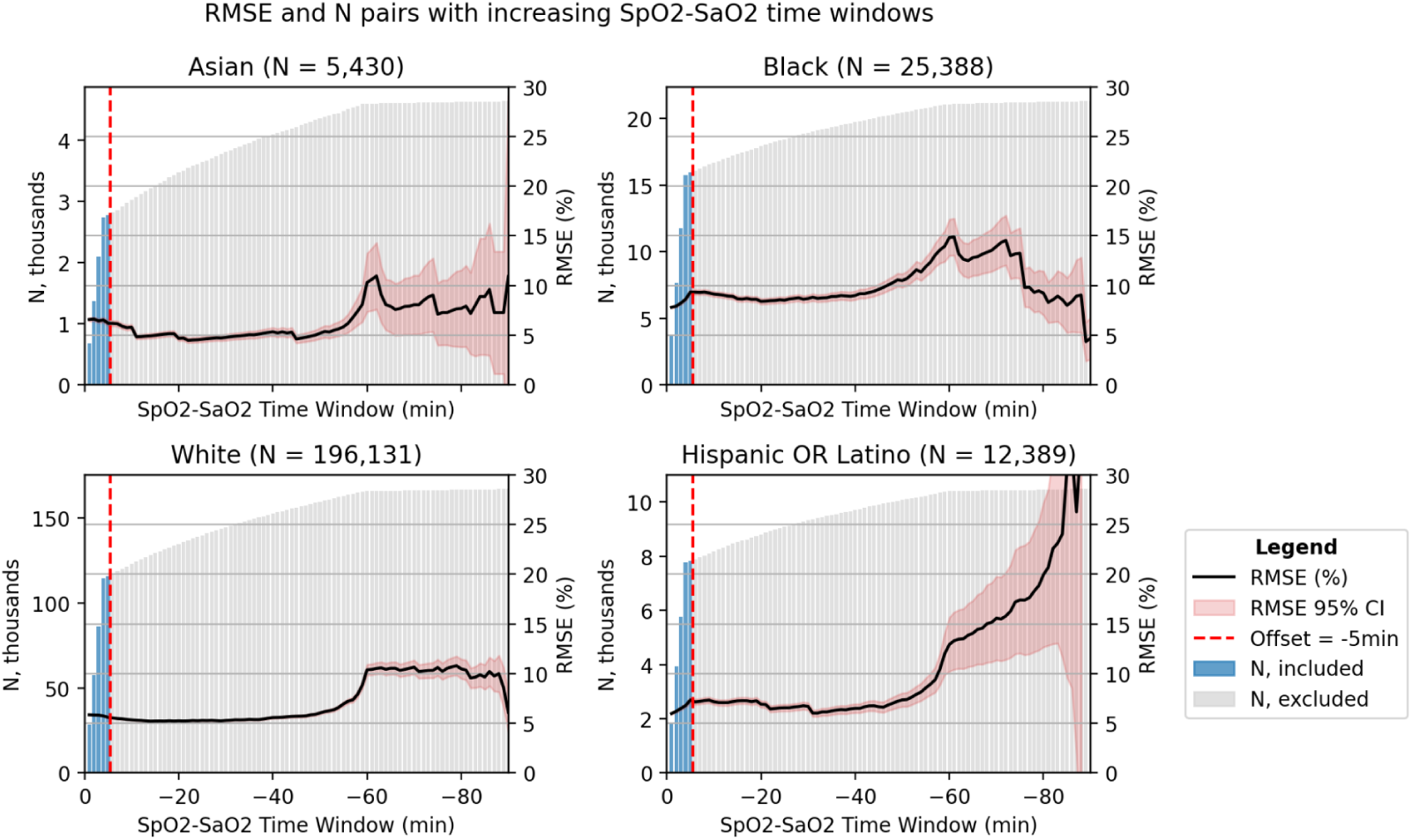
RMSE and number of pairs with varying window between SaO_2_ and SpO_2_, per race and ethnicity, on the extended dataset.

The final dataset consisted of 49,099 first (SaO_2_, SpO_2_) pairs. Most pairs emanated from eICU-CRD (43,438), followed by MIMIC-IV (4,921), and then by MIMIC-III (740) (see figures 3a, 3b, and 3c with the flow diagram per database; figure 4 with the overall flow diagram). The distribution of eligible pairs by race and ethnicity varied across the three databases. Notably, the application of our exclusion criteria in the two MIMIC databases resulted in an overrepresentation of White and male patients, while patients from racial and ethnic groups were dropped at a higher rate. This disproportionate exclusion rate may owe to the lower likelihood of ABG draws among patients from minority racial and ethnic groups.^2^

In sum, across all three databases, White patients formed the most prevalent racial and ethnic subgroup, accounting for ∼ 75% of cases in the resulting dataset after application of our inclusion and exclusion criteria. In addition, the majority of patients in this dataset were male (55.3% in eICU-CRD; 61.6% in MIMIC-III; 65.0% in MIMIC-IV).

As noted in Table 3, the distribution of patients among different regions of the US varied by database, with the Midwest (%), South (%), and West (%) being the primary regions represented overall. The median admission age was approximately 66.0 years in eICU-CRD and 68.0 years in the two MIMIC databases, respectively. The median admission weight, height, and BMI were consistent across the three databases. The median Charlson comorbidity scores were consistent among MIMIC-IV and eICU-CRD; in MIMIC-III, this score was not available. Length of stay (LoS) measures were more variable across databases; in particular, all forms of LoS were consistently shorter in eICU-CRD than in MIMIC-III and MIMIC-IV (p < .001, as determined by a Kruskal-Wallis test). In-hospital mortality rates were found to be 17.8% in eICU-CRD, 17.4% in MIMIC-III, and 15.5% in MIMIC-IV (p < .001, as determined by a Chi-squared test). These differences reflect significant differences across healthcare systems and should be handled carefully when using BOLD.

**Table 3.**
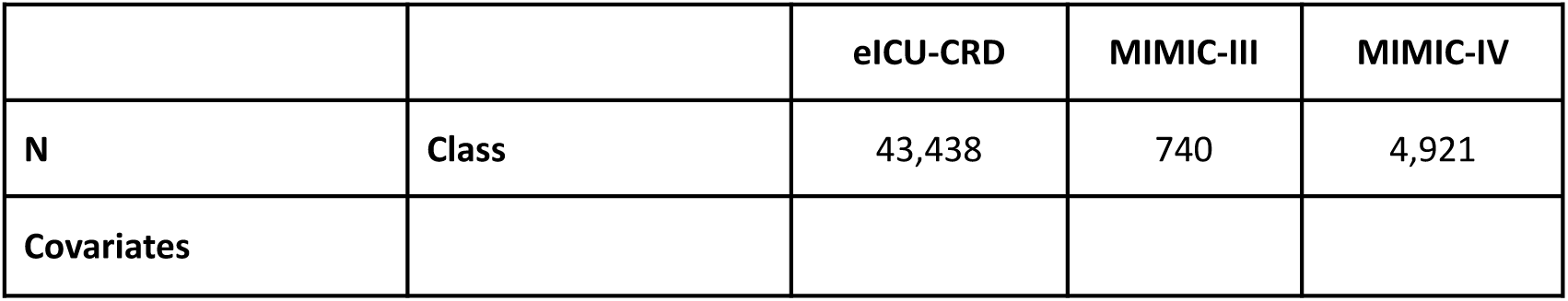
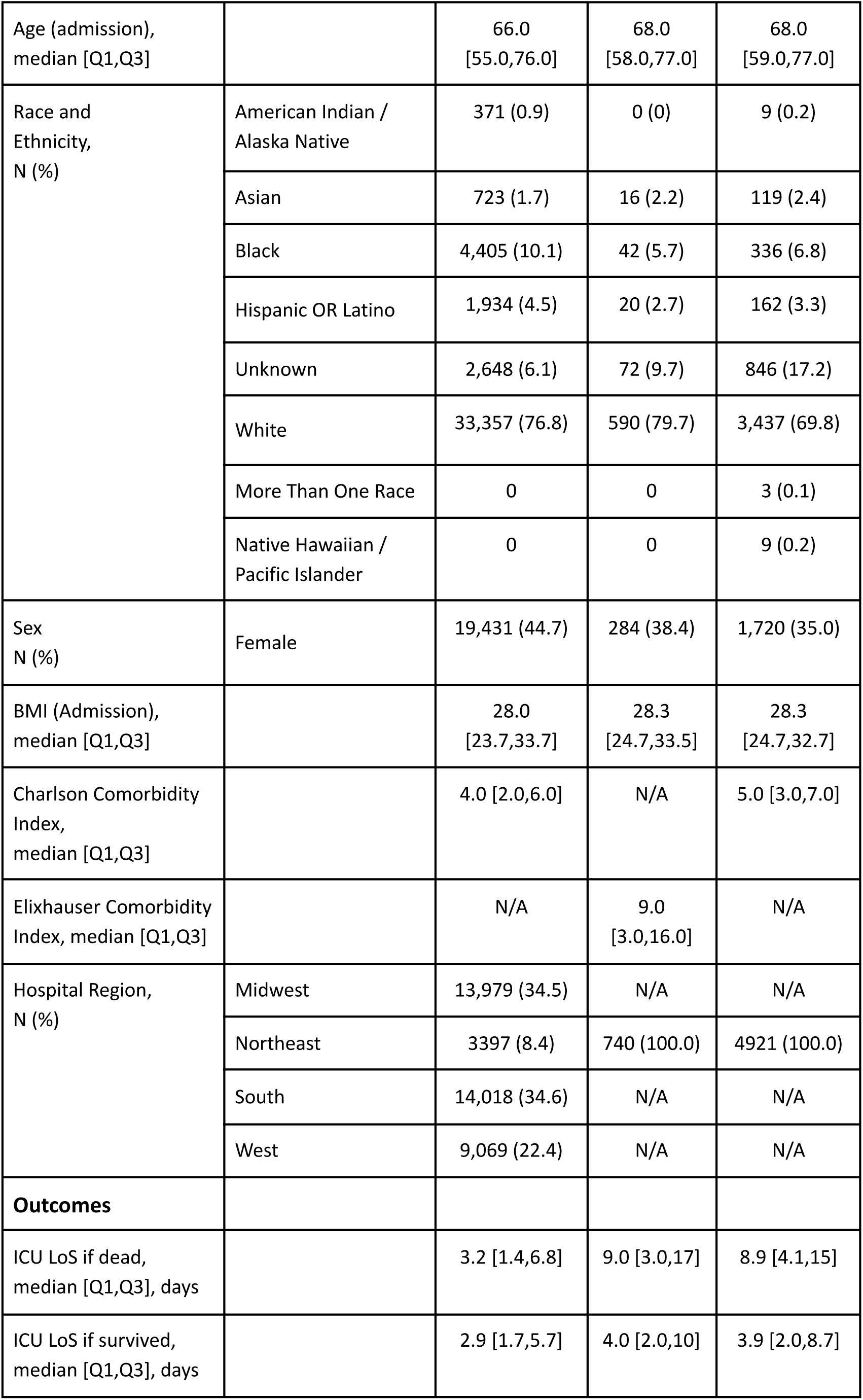
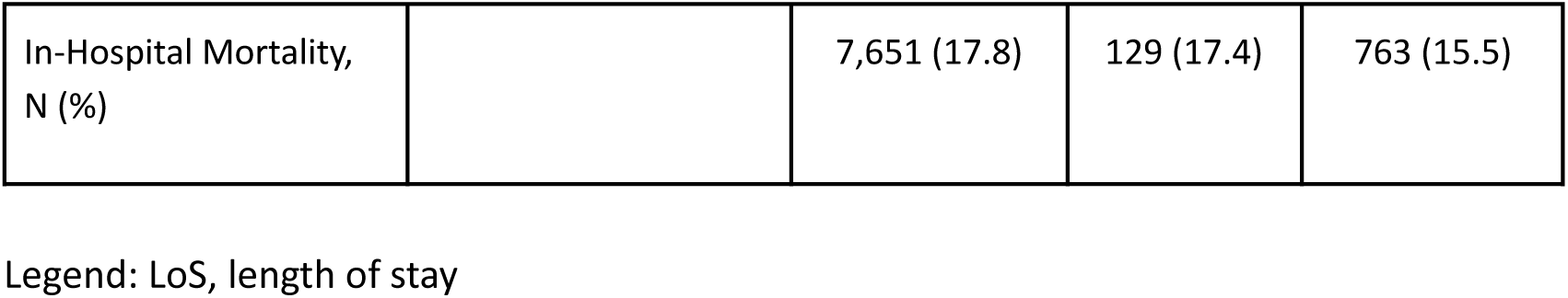
Descriptive patient characteristics by individual dataset.

Static variables, such as the patient’s biological sex, or outcomes, like in-hospital mortality, had very few missing values (see Figure 8 for covariates’ completeness). However, time-varying variables, such as laboratory test values, were more sporadic. For lab tests with significant temporal volatility (e.g., lactate), data up to a maximum of 4 hours before the SaO_2_ measurement were considered. For lab tests often drawn on a daily basis (e.g., basic metabolic panel), this window was extended to 24 hours before baseline. For labs drawn less frequently (e.g., hepatic function panel/complete metabolic panel), data up to 7 days before baseline were included.

**Figure 8.**
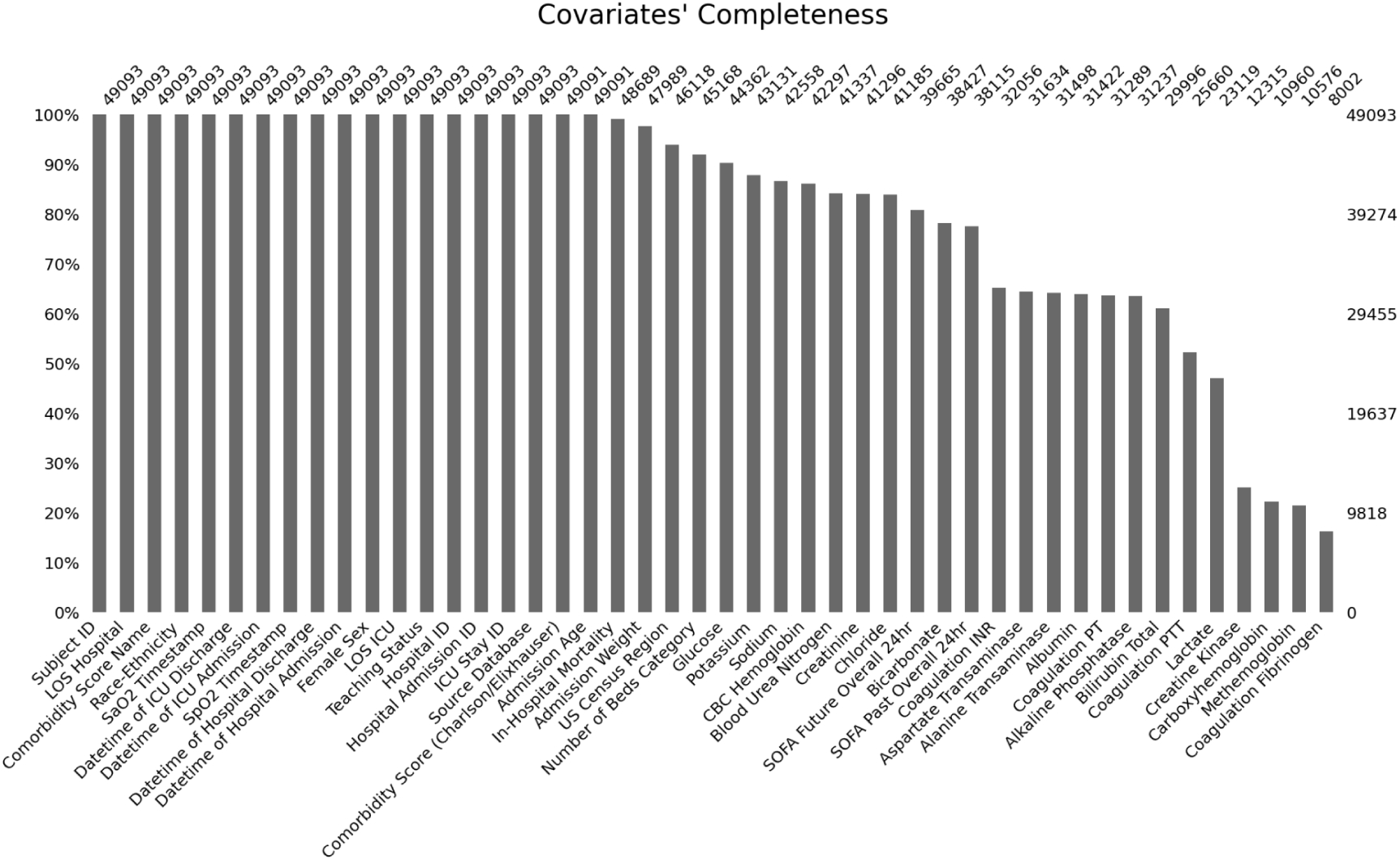
Completeness of the aligned covariates in the preprocessed dataset.

Researchers should carefully choose the most appropriate time window length for their study, based on our data. If they choose to include repeated measurements of the (SaO_2_, SpO_2_) pair for the same patient in their analysis, we strongly recommend replicating our data validation steps to mitigate the risk of introducing systematic errors and limit selection bias. To ensure the highest level of data fidelity, we suggest that a cross-disciplinary team of data scientists and domain experts be involved in the data analysis process.

#### Limitations

There are several noteworthy limitations associated with the preprocessed dataset presented in this paper that researchers and clinicians should consider.

First, imbalances in the sampling rate of arterial blood gas (ABG) across patient sociodemographics, including by race and ethnicity, limit the potential for downstream model development ^2^. Indeed, low ABG sampling rates make it challenging to merge pulse oximeter readings with gold-standard ABG data effectively to characterize outcome heterogeneity in the population and in sufficient quantity to train correction models.^2^ For example, the absence of a uniform rate of ABG sampling across sociodemographics may result in the poor estimation of the differential prevalence of hidden hypoxemia in subpopulations, thereby hindering the evaluation of disparities and the downstream implementation of subpopulation-specific corrections. This issue adds to the reality of limited patient sample sizes for certain racial and ethnic subgroups, posing a further challenge for recalibration efforts aimed at addressing documented disparities and those yet to be identified.

Second, a more general limitation of EHR data that affects our preprocessed dataset is the lack of objective information on skin tone, a factor known to bias pulse oximeter readings. We advocate for efforts to encourage the recording of such variables upon hospital entry in future EHR systems, in order to better address disparities.

#### Strengths

Despite its limitations, our dataset offers several strengths that make it a robust foundation for future research. Notably, it provides a unique platform for quantifying the extent racial and ethnic disparities in intensive care, thereby laying the groundwork for innovative, data-driven solutions to enhance the outcomes of critically-ill patients.

We contribute almost fifty thousand rigorously paired (SaO_2_, SpO_2_) measurements, obtained under strict, clinically relevant criteria. Our curated dataset eliminates a key barrier to entry in the field of data science for critical care. Its creation required the involvement of specialized and multidisciplinary teams of data scientists and clinicians who can navigate complex EHR databases from different health systems. With its public release, our hope is that it will serve as a test bed for future generations of trainees.

The data formats we present are harmonized, user-friendly, and well documented. Original identifiers are kept in the curated dataset to allow the inclusion of further information from the original MIMIC and eICU databases by future users, upon its release in open access.

Finally, to facilitate broader use and encourage careful data engineering, we present what we believe to be a set of best practices in the field of data curation for health equity research. Our methodology for curating EHR data — specifically arterial blood gas and pulse oximetry readings — is fully accompanied by open-source code, which can be easily modified by interested users to accommodate new needs.

### Usage Notes

The data of this paper employs three publicly available datasets MIMIC-III, MIMIC-IV, and eICU-CRD, all available on PhysioNet as CSV files, or on Google Cloud BigQuery.

BOLD is available on PhysioNet as a credentialled database. To access BOLD, users must be registered on PhysioNet, have proper ethics training, and sign a data use agreement outlining the data usage and security standards, prohibiting any effort to identify the patients of the dataset.

We also share on GitHub all the code to recreate the dataset curation process. https://github.com/joamats/pulse-ox-dataset

The 1_dataset.ipynb notebook contains all the necessary queries optimized to be used on Google’s BigQuery (SQL standard) to generate the final CSV file. We did softcode the important inclusion criteria of lower SaO_2_, upper SaO_2_, and lower and upper time windows to facilitate any changes to these key parameters. Analysts need to make sure they set up a BigQuery project according to the instructions in our notebook. We also share the notebooks 2_CONSORT_diagram.ipynb, 3_tableones.ipynb, and 4_technical_validation.ipynb to recreate all the analyses provided in this paper.

https://docs.google.com/spreadsheets/d/1W4PS3-jF3m8OemERsv2r_b9sfACWIr-JQcPxW2A7

This Google’s Spreadsheet file contains the details for all time-varying variables that are encoded, as well as the necessary field for them to be pulled from the databases. These details can be changed either globally, or separately for each database.

https://docs.google.com/spreadsheets/d/1Hv_sOd0--6TPYiB3Crjdn_JrIhIazXXJc05mL4GefOU

Finally, this Google’s Spreadsheet file contains the unified mappings for the static variables (for reference), as well as the race and ethnicity mappings (which are then fed to the created scripts).

## Code Availability

All code used for data extraction, processing, visualization, and technical validation is available as SQL queries (Google’s Bigquery syntax) and Jupyter notebooks in the corresponding PhysioNet page and on GitHub.

https://github.com/joamats/pulse-ox-dataset

The publicly-available scripts are structured as follows:

1. The folders MIMIC-III, MIMIC-IV, and eICU-CRD contain the SQL queries to fetch the data, alongside auxiliary tables that need to be created first in a user’s BigQuery environment.
2. The source folder contains the Jupyter notebook (1_dataset.ipynb) to create the dataset, which is calling the main SQL scripts needed to create the final CSV file. It also contains the notebooks *2_CONSORT_diagram.ipynb*, *3_tableones.ipynb*, and *4_technical_validation.ipynb* to recreate all the analyses.

## Data Availability

The created dataset is under-review on PhysioNet and will be published shortly.
The data of this paper employs three publicly available datasets MIMIC-III, MIMIC-IV, and
eICU-CRD, all available on PhysioNet.

https://physionet.org/content/mimiciv/2.2/

https://physionet.org/content/eicu-crd/2.0/

https://physionet.org/content/mimiciii/1.4/

## Acknowledgements

We would like to thank Tom Pollard for providing thoughtful and constructive suggestions for improving the dataset.

## Author contributions

All authors contributed to writing the manuscript. JM, TS, and AIW collaborated on the data extraction, visualization, and analysis. JM, TS, JG, LN, MLC, XL, JSC, NEZ, KSJ, NB, and JG interpreted, validated results, design of the work and supervised data extraction. LAC and AIW reviewed the paper and supervised the work.

## Supplemental Tables

**Supplemental Table 1.**
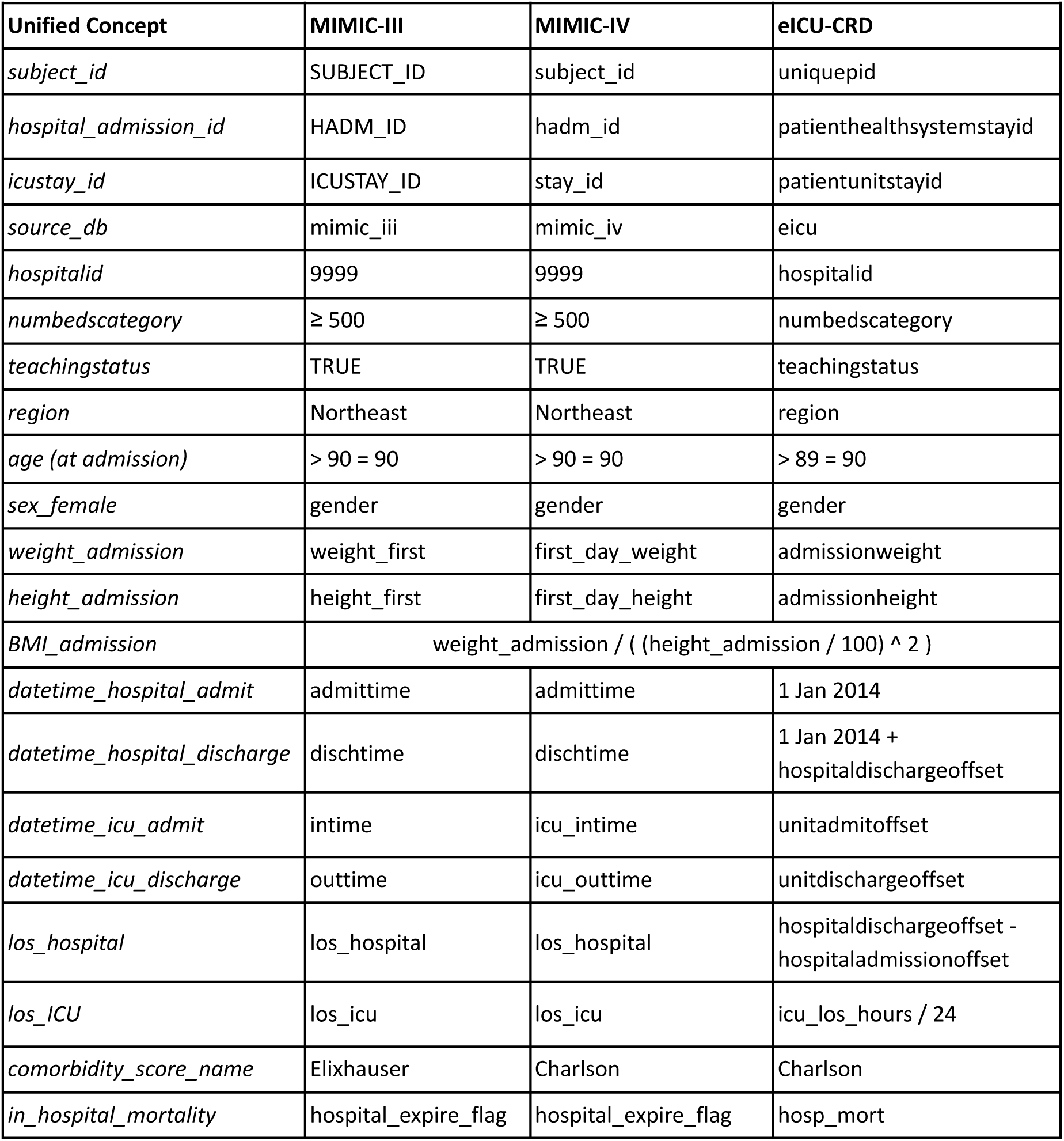
Mapping of MIMIC-III, MIMIC-IV, eICU-CRD static concepts.

**Supplemental Table 2a.** race and ethnicity unified mapping in MIMIC-IV.

| Original Labels in MIMIC-IV | N | Unified Mapping |
| --- | --- | --- |
| AMERICAN INDIAN/ALASKA NATIVE | 140 | American Indian / Alaska Native |
| ASIAN | 840 | Asian |
| ASIAN - ASIAN INDIAN | 179 | Asian |
| ASIAN - CHINESE | 792 | Asian |
| ASIAN - KOREAN | 51 | Asian |
| ASIAN - SOUTH EAST ASIAN | 293 | Asian |
| BLACK/AFRICAN | 309 | Black |
| BLACK/AFRICAN AMERICAN | 6723 | Black |
| BLACK/CAPE VERDEAN | 502 | Black |
| BLACK/CARIBBEAN ISLAND | 426 | Black |
| HISPANIC OR LATINO | 724 | Hispanic OR Latino |
| HISPANIC/LATINO - CENTRAL AMERICAN | 47 | Hispanic OR Latino |
| HISPANIC/LATINO - COLUMBIAN | 60 | Hispanic OR Latino |
| HISPANIC/LATINO - CUBAN | 72 | Hispanic OR Latino |
| HISPANIC/LATINO - DOMINICAN | 535 | Hispanic OR Latino |
| HISPANIC/LATINO - GUATEMALAN | 168 | Hispanic OR Latino |
| HISPANIC/LATINO - HONDURAN | 61 | Hispanic OR Latino |
| HISPANIC/LATINO - MEXICAN | 68 | Hispanic OR Latino |
| HISPANIC/LATINO - PUERTO RICAN | 902 | Hispanic OR Latino |
| HISPANIC/LATINO - SALVADORAN | 104 | Hispanic OR Latino |
| MULTIPLE RACE/ETHNICITY | 68 | More Than One Race |
| NATIVE HAWAIIAN OR OTHER PACIFIC ISLANDER | 110 | Native Hawaiian / Pacific Islander |
| OTHER | 2368 | Unknown |
| PATIENT DECLINED TO ANSWER | 425 | Unknown |
| PORTUGUESE | 322 | White |
| SOUTH AMERICAN | 64 | Hispanic OR Latino |
| UNABLE TO OBTAIN | 844 | Unknown |
| UNKNOWN | 6415 | Unknown |
| WHITE | 47197 | White |
| WHITE - BRAZILIAN | 147 | White |
| WHITE - EASTERN EUROPEAN | 164 | White |
| WHITE - OTHER EUROPEAN | 1307 | White |
| WHITE - RUSSIAN | 754 | White |

**Supplemental Table 2b.** race and ethnicity unified mapping in MIMIC-III.

| Original Labels in MIMIC-III | N | Unified Mapping |
| --- | --- | --- |
| AMERICAN INDIAN/ALASKA NATIVE | 54 | American Indian / Alaska Native |
| AMERICAN INDIAN/ALASKA NATIVE FEDERALLY RECOGNIZED TRIBE | 3 | American Indian / Alaska Native |
| ASIAN | 1510 | Asian |
| ASIAN - ASIAN INDIAN | 93 | Asian |
| ASIAN - CAMBODIAN | 22 | Asian |
| ASIAN - CHINESE | 278 | Asian |
| ASIAN - FILIPINO | 27 | Asian |
| ASIAN - JAPANESE | 7 | Asian |
| ASIAN - KOREAN | 12 | Asian |
| ASIAN - OTHER | 18 | Asian |
| ASIAN - THAI | 4 | Asian |
| ASIAN - VIETNAMESE | 53 | Asian |
| BLACK/AFRICAN | 44 | Black |
| BLACK/AFRICAN AMERICAN | 5591 | Black |
| BLACK/CAPE VERDEAN | 206 | Black |
| BLACK/HAITIAN | 105 | Black |
| CARIBBEAN ISLAND | 9 | Hispanic OR Latino |
| HISPANIC OR LATINO | 1742 | Hispanic OR Latino |
| HISPANIC/LATINO - CENTRAL AMERICAN (OTHER) | 13 | Hispanic OR Latino |
| HISPANIC/LATINO - COLOMBIAN | 10 | Hispanic OR Latino |
| HISPANIC/LATINO - CUBAN | 24 | Hispanic OR Latino |
| HISPANIC/LATINO - DOMINICAN | 83 | Hispanic OR Latino |
| HISPANIC/LATINO - GUATEMALAN | 39 | Hispanic OR Latino |
| HISPANIC/LATINO - HONDURAN | 4 | Hispanic OR Latino |
| HISPANIC/LATINO - MEXICAN | 12 | Hispanic OR Latino |
| HISPANIC/LATINO - PUERTO RICAN | 237 | Hispanic OR Latino |
| HISPANIC/LATINO - SALVADORAN | 18 | Hispanic OR Latino |
| MIDDLE EASTERN | 44 | White |
| MULTI RACE ETHNICITY | 137 | More Than One Race |
| NATIVE HAWAIIAN OR OTHER PACIFIC ISLANDER | 18 | Native Hawaiian / Pacific Islander |
| OTHER | 1549 | Unknown |
| PATIENT DECLINED TO ANSWER | 567 | Unknown |
| PORTUGUESE | 70 | White |
| SOUTH AMERICAN | 9 | Hispanic OR Latino |
| UNABLE TO OBTAIN | 882 | Unknown |
| UNKNOWN/NOT SPECIFIED | 4724 | Unknown |
| WHITE | 42488 | White |
| WHITE - BRAZILIAN | 64 | White |
| WHITE - EASTERN EUROPEAN | 28 | White |
| WHITE - OTHER EUROPEAN | 85 | White |
| WHITE - RUSSIAN | 168 | White |

**Supplemental Table 2b.** race and ethnicity unified mapping in eICU-CRD.

| eICU-CRD | N | Unified Mapping |
| --- | --- | --- |
| — | 2,290 | Unknown |
| African American | 21,308 | Black |
| Asian | 3,270 | Asian |
| Caucasian | 155,285 | White |
| Hispanic | 7,464 | Hispanic OR Latino |
| Native American | 1,700 | American Indian / Alaska Native |
| Other/Unknown | 9,542 | Unknown |

**Supplemental Table 3.**
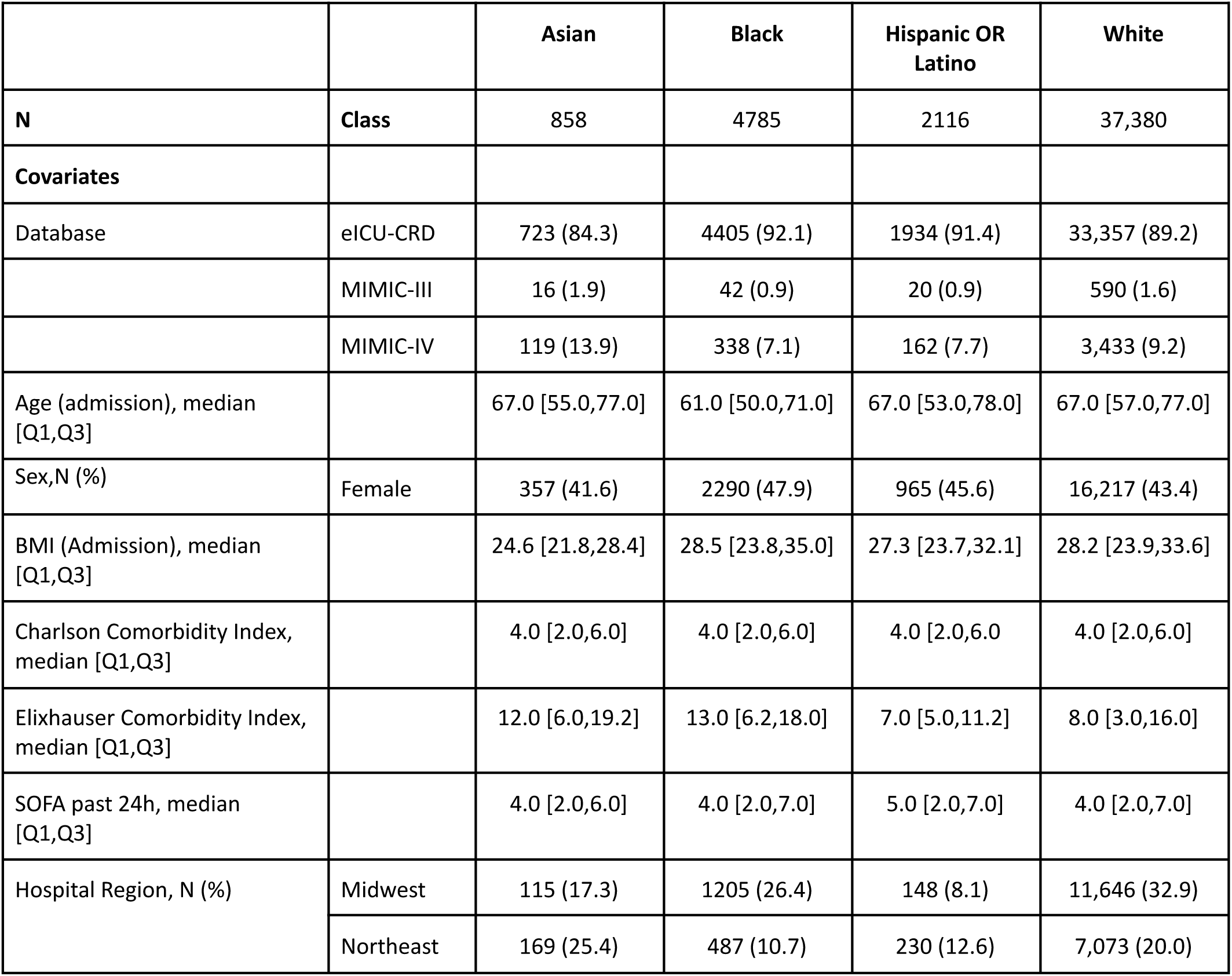
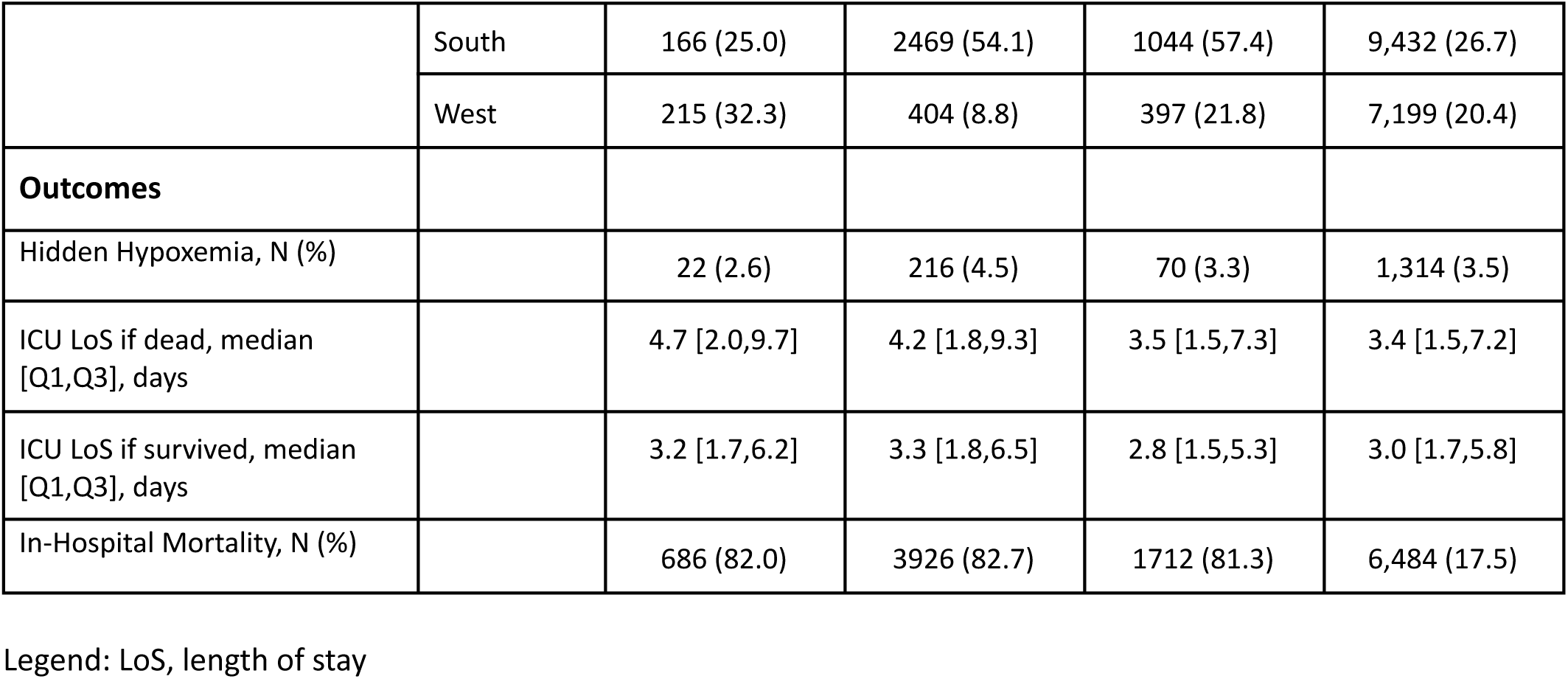
Descriptive patient characteristics by race and ethnicity.

|  |  | Asian | Black | Hispanic OR Latino | White |
| --- | --- | --- | --- | --- | --- |
| **N** | **Class** | 858 | 4785 | 2116 | 37,380 |
| **Covariates** |  |  |  |  |  |
| Database | eICU-CRD | 723 (84.3) | 4405 (92.1) | 1934 (91.4) | 33,357 (89.2) |
|  | MIMIC-III | 16 (1.9) | 42 (0.9) | 20 (0.9) | 590 (1.6) |
|  | MIMIC-IV | 119 (13.9) | 338 (7.1) | 162 (7.7) | 3,433 (9.2) |
| Age (admission), median [Q1,Q3] |  | 67.0 [55.0,77.0] | 61.0 [50.0,71.0] | 67.0 [53.0,78.0] | 67.0 [57.0,77.0] |
| Sex,N (%) | Female | 357 (41.6) | 2290 (47.9) | 965 (45.6) | 16,217 (43.4) |
| BMI (Admission), median [Q1,Q3] |  | 24.6 [21.8,28.4] | 28.5 [23.8,35.0] | 27.3 [23.7,32.1] | 28.2 [23.9,33.6] |
| Charlson Comorbidity Index, median [Q1,Q3] |  | 4.0 [2.0,6.0] | 4.0 [2.0,6.0] | 4.0 [2.0,6.0] | 4.0 [2.0,6.0] |
| Elixhauser Comorbidity Index, median [Q1,Q3] |  | 12.0 [6.0,19.2] | 13.0 [6.2,18.0] | 7.0 [5.0,11.2] | 8.0 [3.0,16.0] |
| SOFA past 24h, median [Q1,Q3] |  | 4.0 [2.0,6.0] | 4.0 [2.0,7.0] | 5.0 [2.0,7.0] | 4.0 [2.0,7.0] |
| Hospital Region, N (%) | Midwest | 115 (17.3) | 1205 (26.4) | 148 (8.1) | 11,646 (32.9) |
|  | Northeast | 169 (25.4) | 487 (10.7) | 230 (12.6) | 7,073 (20.0) |
|  | South | 166 (25.0) | 2469 (54.1) | 1044 (57.4) | 9,432 (26.7) |
|  | West | 215 (32.3) | 404 (8.8) | 397 (21.8) | 7,199 (20.4) |
| **Outcomes** |  |  |  |  |  |
| Hidden Hypoxemia, N (%) |  | 22 (2.6) | 216 (4.5) | 70 (3.3) | 1,314 (3.5) |
| ICU LoS if dead, median [Q1,Q3], days |  | 4.7 [2.0,9.7] | 4.2 [1.8,9.3] | 3.5 [1.5,7.3] | 3.4 [1.5,7.2] |
| ICU LoS if survived, median [Q1,Q3], days |  | 3.2 [1.7,6.2] | 3.3 [1.8,6.5] | 2.8 [1.5,5.3] | 3.0 [1.7,5.8] |
| In-Hospital Mortality, N (%) |  | 686 (82.0) | 3926 (82.7) | 1712 (81.3) | 6,484 (17.5) |

Legend: LoS, length of stay

**Supplemental Table 4.** Descriptive patient characteristics by hidden hypoxemia (SpO_2_ ≥ 88% but SaO_2_ < 88%, as defined by Wong et al.^2^).

|  |  | **Hidden Hypoxemia absent** | **Hidden Hypoxemia present** |
| --- | --- | --- | --- |
| **N** | **Class** | 47,362 | 1,731 |
| **Covariates** |  |  |  |
| Database | eICU-CRD | 41,752 (88.2) | 1,686 (97.4) |
|  | MIMIC-III | 722 (1.5) | 18 (1.0) |
|  | MIMIC-IV | 4,888 (10.3) | 27 (1.6) |
| Age (admission), median [Q1,Q3] |  | 66.0 [55.0,76.0] | 65.0 [54.0,75.0] |
| Race and Ethnicity, N (%) | American Indian / Alaska Native | 371 (0.8) | 9 (0.5) |
|  | Asian | 836 (1.8) | 22 (1.3) |
|  | Black | 4,569 (9.6) | 216 (12.5) |
|  | Hispanic OR Latino | 2,046 (4.3) | 70 (4.0) |
|  | Unknown | 3,462 (7.3) | 100 (5.8) |
|  | White | 36,066 (76.1) | 1,314 (75.9) |
|  | More Than One Race | 3 (0.0) |  |
|  | Native Hawaiian / Pacific Islander | 9 (0.0) |  |
| Sex, N (%) | Female | 20,634 (43.6) | 799 (46.2) |
| BMI (Admission), median [Q1,Q3] |  | 28.1 [23.8,33.5] | 28.2 [23.8,35.6] |
| Charlson Comorbidity Index, median [Q1,Q3] |  | 4.0 [2.0,6.0] | 4.0 [2.0,6.0] |
| Elixhauser Comorbidity Index, median [Q1,Q3] |  | 9.0 [3.0,16.0] | 14.0 [7.2,18.5] |
| SOFA over past 24 hours, median [Q1,Q3] |  | 4.0 [2.0,7.0] | 4.0 [2.0,7.0] |
| Hospital Region, N (%) | Midwest | 13,334 (30.0) | 645 (39.0) |
|  | Northeast | 8,802 (19.8) | 250 (15.1) |
|  | South | 13,564 (30.5) | 454 (27.4) |
|  | West | 8,764 (19.7) | 305 (18.4) |
| **Outcomes** |  |  |  |
| ICU LoS if dead, median [Q1,Q3], days |  | 3.6 [1.6,7.7] | 2.6 [1.0,5.9] |
| LoS ICU if survived, median [Q1,Q3], days |  | 3.0 [1.7,5.9] | 3.6 [1.9,7.0] |
| In-Hospital Mortality, N (%) |  | 8,092 (17.2) | 450 (26.1) |

